# Mapping of SARS-CoV-2 Brain Invasion and Histopathology in COVID-19 Disease

**DOI:** 10.1101/2021.02.15.21251511

**Authors:** Geidy E. Serrano, Jessica E. Walker, Richard Arce, Michael J. Glass, Daisy Vargas, Lucia I. Sue, Anthony J. Intorcia, Courtney M. Nelson, Javon Oliver, Jaclyn Papa, Aryck Russell, Katsuko E. Suszczewicz, Claryssa I. Borja, Christine Belden, Danielle Goldfarb, David Shprecher, Alireza Atri, Charles H. Adler, Holly A. Shill, Erika Driver-Dunckley, Shyamal H. Mehta, Benjamin Readhead, Matthew J. Huentelman, Joseph L. Peters, Ellie Alevritis, Christian Bimi, Joseph P. Mizgerd, Eric M. Reiman, Thomas J. Montine, Marc Desforges, James L. Zehnder, Malaya K. Sahoo, Haiyu Zhang, Daniel Solis, Benjamin A. Pinsky, Michael Deture, Dennis W. Dickson, Thomas G. Beach

**Affiliations:** Banner Sun Health Research Institute, Sun City, AZ; Brigham and Women’s Hospital and Harvard Medical School, Boston, MA; Mayo Clinic College of Medicine, Mayo Clinic Arizona, Scottsdale, AZ; Barrow Neurological Institute, Phoenix, AZ; Arizona State University-Banner Neurodegenerative Disease Research Center, Tempe, AZ; Translational Genomics Research Institute, Neurogenomics Division, Phoenix, AZ; Banner Boswell Medical Center, Sun City, AZ; Banner University Medical Center, Tucson, AZ; Pulmonary Center, Boston University School of Medicine, Boston, MA; Banner Alzheimer’s Institute, Phoenix, AZ; Stanford University School of Medicine, Department of Pathology, Stanford, CA; Centre Hospitalier Universitaire Sainte-Justine, Laboratory of Virology, Montreal, Canada; Stanford University Department of Medicine, Division of Infectious Diseases and Geographic Medicine, Stanford, CA; Mayo Clinic College of Medicine, Mayo Clinic Florida, Jacksonville FL

**Keywords:** coronavirus, neuropathology, RT-PCR, encephalitis, infarction, hypoxia, hemorrhage

## Abstract

The coronavirus SARS-CoV-2 (SCV2) causes acute respiratory distress, termed COVID-19 disease, with substantial morbidity and mortality. As SCV2 is related to previously-studied coronaviruses that have been shown to have the capability for brain invasion, it seems likely that SCV2 may be able to do so as well. To date, although there have been many clinical and autopsy-based reports that describe a broad range of SCV2-associated neurological conditions, it is unclear what fraction of these have been due to direct CNS invasion versus indirect effects caused by systemic reactions to critical illness. Still critically lacking is a comprehensive tissue-based survey of the CNS presence and specific neuropathology of SCV2 in humans. We conducted an extensive neuroanatomical survey of RT-PCR-detected SCV2 in 16 brain regions from 20 subjects who died of COVID-19 disease. Targeted areas were those with cranial nerve nuclei, including the olfactory bulb, medullary dorsal motor nucleus of the vagus nerve and the pontine trigeminal nerve nuclei, as well as areas possibly exposed to hematogenous entry, including the choroid plexus, leptomeninges, median eminence of the hypothalamus and area postrema of the medulla. Subjects ranged in age from 38 to 97 (mean 77) with 9 females and 11 males. Most subjects had typical age-related neuropathological findings. Two subjects had severe neuropathology, one with a large acute cerebral infarction and one with hemorrhagic encephalitis, that was unequivocally related to their COVID-19 disease while most of the 18 other subjects had non-specific histopathology including focal β-amyloid precursor protein white matter immunoreactivity and sparse perivascular mononuclear cell cuffing. Four subjects (20%) had SCV2 RNA in one or more brain regions including the olfactory bulb, amygdala, entorhinal area, temporal and frontal neocortex, dorsal medulla and leptomeninges. The subject with encephalitis was SCV2-positive in a histopathologically-affected area, the entorhinal cortex, while the subject with the large acute cerebral infarct was SCV2-negative in all brain regions. Like other human coronaviruses, SCV2 can inflict acute neuropathology in susceptible patients. Much remains to be understood, including what viral and host factors influence SCV2 brain invasion and whether it is cleared from the brain subsequent to the acute illness.

## INTRODUCTION

The coronavirus SARS-CoV-2 (SCV2) emerged in Wuhan, China, in late 2019 and then rapidly spread worldwide. It causes severe acute respiratory distress with frequent systemic complications, termed COVID-19 disease, with substantial morbidity and mortality. As SCV2 shares RNA sequences with two previously studied coronaviruses, SARS-CoV and MERS-CoV, that caused similar human syndromes, and as there is evidence from both human autopsies and animal models that both of these may invade the brain and cause neurological symptoms ^1–11^, it seems likely that SCV2 may have similar capabilities. The human coronavirus OC43 has caused acute encephalomyelitis in children ^12–14^, other coronaviruses have been associated with chronic demyelinating disease in animal models ^15–20^, and there have been suggestions of an association with human multiple sclerosis ^21^. The mouse hepatitis virus MHV-A59, a coronavirus, and JHM strains of murine coronaviruses are neurotropic and virulent in rodents ^22,23^ and nonhuman primates ^24–27^. Two human coronaviruses, strains 229E and OC43, were detected by RT-PCR, Northern hybridization and in-situ hybridization in 44% and 23%, respectively, of 90 human brains obtained from multiple brain banks throughout Europe, the UK and USA ^21^, suggesting that coronaviruses are capable of CNS invasion and persistence.

Although there have been many clinical reports and reviews that collectively describe a broad range of neurological signs, symptoms and syndromes that have occurred in humans infected with SCV2 ^28–35^, it is still unclear what fraction of these have been due to direct viral brain invasion versus indirect structural or functional changes caused by systemic reactions to critical illness, including coagulopathy, sepsis, autoimmune mechanisms or multiorgan failure ^36^. The majority of neurological changes have not been severe but between 2 and 6% of clinically-affected subjects have had acute stroke, and there has been a smaller percentage reported with encephalitis or Guillain-Barré syndrome. The subacute and long-term neurological consequences have not been sufficiently studied. Still critically lacking is a comprehensive tissue-based survey of the CNS presence or specific neuropathology of SCV2 in humans.

Autopsy reports to date have described meningitis and/or encephalitis in small numbers of patients dying with COVID-19 disease ^37–45^, and more frequently, both ischemic and/or hemorrhagic acute or subacute cerebrovascular lesions ^37,39,44,46–57^, but these have not always been supported by direct evidence of CNS SCV2 presence. Elements of the classical neuropathology of viral CNS infections ^58,59^, including lymphocytic leptomeningitis and encephalitis, microglial nodules, perivascular lymphocytic cuffing, focal demyelination and viral inclusions, have most often been absent. There have been at least 16 published studies ^36,38,39,45,47–50,60–67^ that used gold-standard RT-PCR methods to interrogate SCV2 genomic presence in postmortem brain tissue, but each of these have examined relatively few brain regions. The brain has hundreds of anatomically-distinct regions with possibly thousands of different cell phenotypes. In many viral CNS infections there is a pronounced cell selectivity ^58,59,68,69^. Assaying for the presence of SCV2 in only a few brain regions is likely to underestimate the overall CNS prevalence.

With so many different brain regions, deciding which to examine for viral presence is difficult but can be guided by available prior virology. Viruses enter the brain through two major routes. Entry may occur through peripheral endings of the cranial nerves, or through the blood, the latter through either direct transgression of the blood-brain-barrier or by initial colonization of endothelial cells ^58,59,63,70^. For SCV2, a leading candidate for viral brain entry is the nasal olfactory mucosa, with its direct neuronal connection to the olfactory bulb and tract ^71,72^ this is also suggested by reported anosmia in SCV2 ^73,74^ and by strong olfactory mucosal expression of angiotensin converting enzyme-2 (ACE2) and neuropilin-1, possible cellular access cofactors for SCV2 ^4,9,75–78^. From the olfactory bulb there are direct neuronal connections to the amygdala, entorhinal area and hippocampus. Many brain regions and several cell types have been reported to express ACE2 or other proteins implicated in COVID-19 pathogenesis ^36,61,79–81^ but, aside from the olfactory bulb, much attention has been focused on the brainstem as it was reportedly affected in the 2003 SARS-CoV epidemic and in animal models of viral infection, possibly indicative of entry through the cranial nerves and in particular through respiratory and gastrointestinal tract epithelium and thence the vagus nerve ^1,82,83^. Human coronavirus experimental models have shown selective brain localization as well as axonal transport and hematogenous CNS entry ^24,25,82,84–86^.

We sought to address the deficiency of neuroanatomically-detailed SCV2 localization data by comprehensively surveying, with calibrated RT-PCR assays, for the presence of genomic SCV2 RNA in 16 brain regions from 20 subjects who died of COVID-19 disease as well as 4 subjects dying in the same time period without COVID-19. Targeted areas included those with direct connections to the peripheral nervous system through cranial nerves, including the olfactory bulb, medullary dorsal motor nucleus of the vagus nerve, and pontine trigeminal nerve nuclei, as well as areas possibly exposed to hematogenous entry, including the choroid plexus and leptomeninges, which are primarily composed of blood vessels, as well as the median eminence of the hypothalamus and area postrema of the medulla, where the blood-brain-barrier is deficient.

## MATERIALS AND METHODS

### Human Subjects

Subjects received neuropathological examinations at either Banner Sun Health Research Institute or Mayo Clinic Jacksonville.

### Banner Sun Health Research Institute Subjects

The Institute is located in Sun City, Arizona, a suburb of Phoenix. Subjects were enrolled in one of two separate Institutional Review Board (IRB) approved protocols reviewed by the Western IRB in Puyallup, Washington. Five of the ten COVID-19 disease subjects and the 4 non-COVID-19 disease control cases were volunteers who had donated their brains and/or bodies for research as part of the Arizona Study of Aging and Neurodegenerative Disorders ^87^ and Brain and Body Donation Program (AZSAND/BBDP; https://www.brainandbodydonationregistration.org/), a longitudinal clinicopathological study and biospecimen resource for normal aging and age-related diseases. All of these subjects or their legal representatives signed an IRB-approved informed consent form allowing both standardized research clinical assessments during life and several options for brain and/or bodily organ donation after death. Clinical data was also collected from their private medical records. Another 5 subjects were enrolled from hospitals in Tucson or Phoenix, Arizona, immediately before or after death, specifically on the basis of having died with severe COVID-19 disease. Clinical data was collected from their hospitals’ and/or physicians’ office records. For all subjects, the left side of each brain was fixed for four days in 10% formalin at 4°C while the right side was sliced into 1 cm coronal slabs and frozen on dry ice, then stored at −80°C.

### Mayo Clinic Jacksonville

All subjects except M10 had been clinically diagnosed with parkinsonism and/or dementia, and all had died with clinically diagnosed COVID-19 disease (Tables 1, 3 and 4). Their brains were removed at several locations throughout the United States, with both fixed and frozen portions taken, following which these were sent to the Mayo Clinic in Jacksonville, Florida for neuropathological examination and diagnosis.

**Table 1.** Case data for Covid-19 disease-related signs, symptoms, treatment and diagnosis. B = BSHRI AZSAND protocol;; np = nasopharyngeal; am = antemortem; pm = postmortem; C = Caucasian;. Symptom abbreviations: ArtThr = peripheral arterial thrombosis; Diarr = diarrhea; Enceph = encephalopathy; Fev = fever; HA = headache; Ftg = fatigue; Naus = nausea; RespFail = respiratory failure; SOB =shortness of breath; Str = stroke; Sz = seizure-like activity; Vom = vomiting; Treatment abbreviations: Abx = anti-bacterial antibiotics; ACoag = anti-coagulants; ConPlas = convalescent plasma; HF = high-flow; ICU – intensive care unit; LF =low-flow; MechVent = mechanical ventilation; Morph = morphine; NIPPV = non-invasive positive pressure ventilation; O2 = oxygen; Rem = remdesivir. N/A = details not available.

| Case | Age | Sex | Race | Signs/Symptoms | Covid-Related Treatment | SCV2 Dx |
| --- | --- | --- | --- | --- | --- | --- |
| B1 | 90s | M | C | Fev; SOB; Cough; RespFail | Hospice; LFO2 | npPCRpm |
| B2 | 70s | F | C | Fev; SOB; Cough; RespFail | Hospice; LFO2; Morph | npPCRpm |
| B3 | 70s | M | C | Fev; Hypox; Naus; Vom; Diarr; Ftg; SOB; ArtThr; Str; RespFail | ICU; MechVent O2; Dex; Abx; Hep; Acoag; Rem | npPCRam<br>npPCRpm |
| B4 | 80s | M | C | SOB; Cough; RespFail | Hospital/Hospice; HF02; NIPPV; Dex; Abx; Acoag; Rem | npPCRam<br>npPCRpm |
| B5b | 90s | M | C | Fev; SOB; Cough; Ftg; Diarr; RespFail | Hospital/Hospice; HF02; NIPPV; Dex; Abx; Acoag; Rem | npPCRam<br>npPCRpm |
| B6 | 70s | M | C | Fev; SOB; Cough; RespFail | Hospice; Morph | npPCRam<br>npPCRpm |
| B7 | 70s | F | C | Fev; Cough; HA; SOB; RespFail | Hospital/Hospice: HFO2, NIPPV; Abx: Dex; Rem; ConPlas | npPCRam<br>npPCRpm |
| B8 | 80s | M | C | SOB; RespFail | Hospice | npPCRpm |
| B9 | 70s | M | C | Fev; Ftg; SOB; RespFail; Enceph | Hospital; NIPPV; Abx; Morph | npPCRam<br>npPCRpm |
| B10 | 60s | F | C | SOB; RespFail; Fev | ICU; MechVent; Dex; Abx; Acoag | npPCRam<br>npPCRpm |
| B11 | 80s | M | C | N/A | N/A | npPCRpm |
| B12 | 90s | M | C | N/A | N/A | npPCRpm |
| B13 | 90s | F | C | N/A | N/A | npPCRpm |
| B14 | 90s | M | C | N/A | N/A | npPCRpm |
| M1 | 70s | F | N/A | N/A | N/A | N/A |
| M2 | 80s | F | N/A | N/A | N/A | N/A |
| M3 | 70s | F | N/A | N/A | N/A | N/A |
| M4 | 80s | M | N/A | N/A | N/A | N/A |
| M5 | 70s | F | N/A | N/A | N/A | N/A |
| M6 | 60s | M | N/A | N/A | N/A | N/A |
| M7 | 70s | F | N/A | N/A | N/A | N/A |
| M8 | 70s | M | N/A | N/A | N/A | N/A |
| M9 | 80s | M | N/A | N/A | N/A | N/A |
| M10 | 30s | M | N/A | Fev; Sz; Enceph | N/A | N/A |
| M11 | 90s | F | N/A | N/A | N/A | N/A |

**Table 2.**
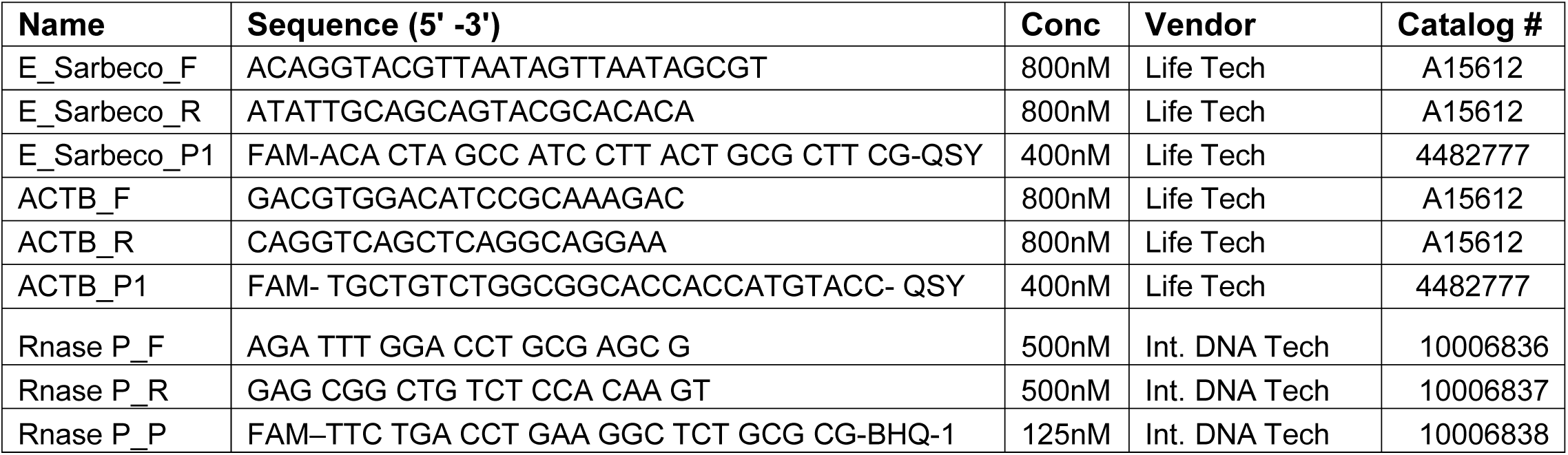
Primers and probes used for RT-PCR. E gene= envelope (*E*) gene; ACTB = Actin Beta; RNASE P= ribonucleus P; F= forward; R= reverse; P= probe; Conc = concentration of primer used; Life Tech. = Life Technologies; Int. DNA Tech. = Integrated DNA Technologies.

**Table 3.** Case data for comorbid conditions. Abbreviations for clinically-diagnosed conditions: Afib = atrial fibrillation; AA = aortic aneurysm; CarD = carotid artery disease; CAD = coronary artery disease; CHF = congestive heart failure; Cig = history of cigarette smoking; Cirrh = hepatic cirrhosis; CKD = chronic kidney disease; CLL = chronic lymphocytic leukemia; COPD = chronic obstructive pulmonary disease; Dem = dementia; DM = diabetes mellitus; DL = dyslipidemia; Emphy = emphysema; Gallst = gallstones; HT = hypertension; MI = myocardial infarction; Ob = obesity; Pace = cardiac pacemaker; Park = parkinsonism; PE = pulmonary embolism; PF = pulmonary fibrosis; PVD = peripheral vascular disease; SLap = sleep apnea; Steat = hepatic steatosis; Str = stroke. Abbreviations for autopsy-diagnosed conditions: AD = Alzheimer’s disease dementia; ARTAG = age-related tau astrogliopathy; ASCVD = atherosclerotic cerebrovascular disease; Crib = Cribriform state; CWMR = cerebral white matter rarefaction; DBS = deep brain stimulator; DLBD = diffuse Lewy body disease; FTLD-TDP = frontotemporal lobar degeneration with TDP-43 proteinopathy; HS = hippocampal sclerosis; Leuko = leukoencephalopathy; LATE = Limbic age-related TDP-43 encephalopathy; MSA = multiple system atrophy; PD = Parkinson’s disease; PART = primary age-related tauopathy; PNLA = pallido-nigro-luysian atrophy; PSP = progressive supranuclear palsy; SC = senile changes (less-than diagnostic plaque and tangle densities); TLBD = transitional Lewy body disease; VaD = vascular dementia. N/A = not available.

| Case | Clinically Diagnosed Conditions | Autopsy Diagnosed Brain Conditions |
| --- | --- | --- |
| B1 | CAD; Cig; COPD; Dem; DM; DL; HT; Ob | AD; VaD; HS; PSP; LATE |
| B2 | Afib; Cig; Dem; HT; PE | PD; AD; ARTAG; DBS |
| B3 | AA; CAD; Cig; COPD; DM; DL; Emphy; Gallst; HT; Ob; PVD; Steat | PART |
| B4 | AA; CAD; CarD; Cig; COPD; DL; Emphy; HT; Ob; PE; PVD; SLap | PART |
| B5 | CAD; Cig; CKD; DM; DL; Emphy; HT; Park; PF | AD; CWMR |
| B6 | CAD; CKD; DL; HT; Ob; Pace; Dem; Park | AD; PSP; TLBD; CWMR; LATE |
| B7 | Afib; Asth; CAD; CHF; CarD; HT; Ob; Pace; SLap; | PART |
| B8 | CAD; Cig; DL; MI | AD; MSA; LATE |
| B9 | DL; DM; Gallst; Ob; SLap; Steat | PART |
| B10 | Cig; Cirrh; DM; Emphy; Gallst; HT | Brain: PART |
| B11 | Afib; CAD; HT; DL; MI; Str; CKD; Cig | N/A |
| B12 | Afib; DL; CLL; CKD; Cig; Ob | N/A |
| B13 | HT; DL; DM; CAD; CKD; COPD; PVD; CRF; Ob | N/A |
| B14 | Afib; HT; PVD; COPD; PE; Str; Cig | N/A |
| M1 | Dem | AD; TLBD; VaD; Leuko; Crib; LATE |
| M2 | Dem | AD; ASCVD; Leuko; Crib |
| M3 | Dem | DLBD; AD |
| M4 | Park | PSP atypical (PNLA); TSA |
| M5 | Dem; Park | PSP; SC; ASCVD; Leuko; Crib |
| M6 | Dem | AD |
| M7 | Dem; Park | FTLD-TDP; VaD; Leuko; SC |
| M8 | Dem; Park | VaD; AD |
| M9 | Dem; Park | PSP (PNLA); PART; ARTAG |
| M10 | N/A | None |
| M11 | N/A | DLB |

**Table 4.** Case data for possible Covid-19 disease-related neuropathological findings. N/A = not applicable. APP = β-amyloid precursor protein.

| **Case** | **Possible Covid-19-Related Neuropathological Findings in the Brain** |
| --- | --- |
| **B1** | Focal leukoencephalopathy with microgliosis; Multifocal sparse perivascular mononuclear cells |
| **B2** | Multifocal sparse perivascular mononuclear cells; Multifocal APP-positive neuronaxonal swellings |
| **B3b** | Acute ischemic & hemorrhagic infarction, middle cerebral artery territory; Multifocal APP-positive neuronaxonal swellings |
| **B4** | Multifocal sparse perivascular mononuclear cells; Multifocal APP-positive neuronaxonal swellings |
| **B5** | Multifocal sparse perivascular mononuclear cells; Multifocal APP-positive neuronaxonal swellings |
| **B6** | Microglial nodules; Multifocal sparse perivascular mononuclear cells |
| **B7** | Multifocal sparse perivascular mononuclear cells; Multifocal APP-positive neuronaxonal swellings |
| **B8** | Acute microscopic infarcts; Multifocal sparse perivascular mononuclear cells; Focal mineralization of pyramidal neurons and multifocal mineralization of blood vessels; Multifocal APP-positive neuronaxonal swellings |
| **B9** | Acute microscopic hemorrhages; Multifocal sparse perivascular mononuclear cells; Multifocal APP-positive neuronaxonal swellings |
| **B10** | Acute microscopic hemorrhage; Sparse to focally marked perivascular mononuclear cells |
| **B11** | N/A (Non-Covid-19 control case) |
| **B12** | N/A (Non-Covid-19 control case) |
| **B13** | N/A (Non-Covid-19 control case) |
| **B14** | N/A (Non-Covid-19 control case) |
| **M1** | None |
| **M2** | Multifocal APP-positive neuronaxonal swellings |
| **M3** | Multifocal APP-positive neuronaxonal swellings |
| **M4** | None |
| **M5** | None |
| **M6** | Multifocal APP-positive neuronaxonal swellings |
| **M7** | None |
| **M8** | Multifocal APP-positive neuronaxonal swellings |
| **M9** | None |
| **M10** | Encephalitis with multifocal sparse perivascular mononuclear cells, acute hemorrhages and uncal herniation; Multifocal APP-positive neuroaxonal swellings |
| **M11** | None |

### Diagnostic Autopsy Examinations

Medically-licensed pathologists performed all examinations. For BSHRI subjects, all but one had whole-body autopsies, with one examination limited to the brain; all 10 Mayo Clinic subject examinations were limited to brain. Those with whole-body exams had extensive lung sampling, bilaterally from upper and lower lobes. Detailed reporting of results for organs other than brain will occur in a separate publication.

Neuropathologists (TGB for BSHRI and DWD for Mayo Clinic) specifically noted the presence or absence of the classical neuropathology of viral CNS infections, including lymphocytic leptomeningitis and encephalitis, microglial nodules, perivascular lymphocytic cuffing, focal demyelination and viral intracellular inclusions. Also noted were any acute or subacute microscopic changes. Otherwise, including for neurodegenerative diseases, the BSHRI neuropathological diagnostic approach has been previously described ^87^. For both BSHRI and Mayo Clinic, published clinicopathological neurodegenerative and cerebrovascular disease consensus criteria ^88–100^ were used when applicable, incorporating research clinical assessment results as well as pertinent private medical history. The histological sampling and staining incorporated the protocols recommended by the National Institute on Aging and Alzheimer’s Association (NIA-AA) ^98–100^.

Immunohistochemical staining for β-amyloid precursor protein (APP) was performed at the Mayo Clinic Florida as previously published ^101^ on sections of anterior cingulate gyrus with corpus callosum and precentral gyrus with adjacent gyral white matter, to assist with the detection of localized axonal swellings indicative of subacute and acute axonal damage, a reported finding in the brains of COVID-19 subjects ^53,56^.

For those BSHRI subjects with whole-body autopsies, formalin-fixed, paraffin-embedded (FFPE) bilateral upper and lower lobe lung samples were stained with hematoxylin and eosin (HE) to assess for the presence of histopathology consistent with COVID-19 pneumonia.

## SARS-CoV-2 RT-PCR methods

### Clinical SCV2 diagnosis

All *in vivo* clinical diagnoses were rendered by licensed medical laboratories using US Food and Drug Administration (FDA) Emergency Use Authorization (EUA) protocols.

### Postmortem SCV2 nasopharyngeal RNA detection

To assess for SCV2 viral presence, postmortem nasopharyngeal swabs (BD Cat # 220531) from all BSHRI cases were assayed for SCV2 RNA using FDA EUA protocols https://www.fda.gov/media/136818/download at Clinical Laboratory Improvement Amendments (CLIA)-approved laboratories, either operated by Sonora Quest, a division of Quest Diagnostics, or by the Stanford Health Care in Stanford, California.

### Postmortem SCV2 RNA detection in blood serum, cerebrospinal fluid, lung and brain tissue

Frozen brain samples were dissected from 16 brain regions, including olfactory bulb, entorhinal area, CA1 region of the hippocampus, amygdala, temporal, frontal and primary visual neocortex, dorsal medulla in the region of the motor nucleus of the vagus nerve and area postrema, pontine tegmentum in the region of the trigeminal nuclei, substantia nigra, hypothalamus in the region of the median eminence, thalamus, putamen at the lentiform nucleus, cerebellar cortex, choroid plexus and leptomeninges. Cases with whole body autopsy were sampled for frozen tissue bilaterally from the upper and lower lung lobes. Aliquots of postmortem intracardiac blood serum and postmortem intracerebroventricular CSF were also assayed.

All RNA isolations and RT-PCR assays were performed at BSHRI. RNA was extracted from 20 mg of frozen brain and lung tissue or 200-250 µl of blood serum and CSF using Qiagen RNeasy Plus Mini Kits (Cat # 74134 for tissue and # 217204 for blood serum and CSF) following the manufacturer’s instructions and eluted in 50 μLof RNAse-free water. SCV2 RNA was detected using previously described primer and probe sequences (Table 2) targeting the envelope (E) gene^102^. RNase P and actin primers and probes were used as housekeeping gene/host tissue amplification controls. RT-PCR was performed in duplicate in separate wells with a 20 µl volume containing 4 μl of RNA and 5 μl of 4X Taqpath One-Step RT-qPCR Master Mix (Cat # A15299 Life Technologies) on Bio-Rad CFX Connect. Inconsistent results, e.g. when one of the duplicates amplified but not the other, were repeated in new duplicate samples.

Thermal cycling was successively performed at 25 °C for 2 minutes for UNG (Uracil N-glycosylase) incubation, 15 minutes at 52 °C for reverse transcription and 2 minutes at 94°C, then consecutively for 15 seconds at 95 °C, 40 seconds at 55 °C and 20 seconds at 68 °C, for a total of 45 cycles.

Preliminary work with primers and probes for the N1 and N2 SCV2 regions, as well as the SCV2 RNA-dependent RNA polymerase (RdRp), had high false-negative rates in lung samples, relative to those for the E gene, and hence these were not used for further assays.

We considered an E gene Ct threshold ≤ 40 as positive. An E gene Ct > 40 with an RNAse P Ct < 35 was considered negative. An E gene Ct > 35 with RNAse P > 35 was considered indeterminate. Positive controls were included in each RT-PCR run; these included a mixture of synthetic SCV2 RNA (RNA transcripts for 5 gene targets, E, N, ORF1ab, RdRP and S genes; Cat # COV019 Bio Rad Labs), as well as a frozen lung sample from a COVID-19 decedent previously shown to be reliably positive. The relative number of E gene copies per µl in brain or lung sample were estimated by interpolation on a standard curve created with serial dilutions of 1:1 synthetic SCV2 RNA (successive dilutions were each 2-fold more dilute).

To evaluate assay validity, RNA aliquots from each of 5 study subjects, with a total of 30 samples, including positive and negative samples of olfactory bulb, dorsal medulla and lung, were analyzed blinded to diagnosis and previous RT-PCR results using similar RT-PCR methods Stanford Health Care in Stanford, California, with complete agreement on positive vs negative results for all anatomical sites and subjects.

## RESULTS

### Clinical Characteristics of Study Subjects

All of the COVID-19 disease cases were Caucasian and had died with clinical diagnoses of respiratory failure (Table 1). Of the non-COVID-19 disease control cases (Table 1, B11-B14), one (B11) had died with pneumonia and sepsis, two (B12, B13) had died of unspecified “natural causes” and one (B14) had died of complications of chronic lymphocytic leukemia. All COVID-19 subjects had one or more pre-existing medical conditions known to be associated with clinically serious COVID-19 disease (Table 3). Three of the BSHRI COVID-19 subjects had been clinically diagnosed with dementia and two with parkinsonism. All but two of the BSHRI COVID-19 subjects had been clinically diagnosed with severe COVID-19 disease, by *in vivo* RT-PCR confirmation of SCV2 presence by nasopharyngeal swab assays; two had RT-PCR diagnosis done at postmortem alone. Six of the BSHRI COVID-19 cases had received typical treatment regimens for COVID-19 disease; four had received only hospice or “comfort” measures. Those BSHRI COVID-19 subjects enrolled through AZSAND were not significantly different in terms of age or sex (age range 72-93, mean 80.8 SD 9.4; one female, 4 males) but had shorter postmortem intervals (PMI; range 2.5-11.6 hours, mean 5.7 SD 3.4) than those enrolled through the COVID-19-dedicated protocol (age range 67-97, mean 78.0 SD 11.6; two females, 4 males; PMI range 3.5-24.9 hours, mean 17.9 SD 8.9). BSHRI non-COVID-19 control subjects were older (age range 87-97, mean 92.5 SD 4.4) and had shorter postmortem intervals (PMI range 1.9-3.5 hours, mean 3.0 SD 0.73) than either of the BSHRI COVID-19 case groups, but sex distribution was similar (three males, one female). All BSHRI subjects had confirmation of SCV2 presence or absence by RT-PCR SCV2 assay of postmortem nasopharyngeal swab as well as by having postmortem lung histology (for the cases with whole-body autopsy) consistent with COVID-19 pneumonia. Eight of nine BSHRI subjects with frozen lung tissue available were positive in lung samples for SCV2 RNA amplification.

Mayo Clinic subjects were significantly younger than the BSHRI combined COVID-19 and non-COVID-19 subjects (Table 1), with more females than males (Mayo Clinic subjects age range 38-97, mean 76.2 SD 14.9, 6 female, 4 male; BSHRI subjects age range 67-97, mean 79.3 SD 10.0; 7 male, 3 female). All had been referred to the Mayo Clinic primarily for neurodegenerative disease diagnosis but all also had clinically-diagnosed severe COVID-19 disease. Seven had been clinically diagnosed with dementia and five with parkinsonism or possible parkinsonism. Details of their COVID-19 diagnosis and treatment, or of their pre-existing general medical comorbidities are not available.

### Neuropathological Findings in Study Subjects

All COVID-19 disease subjects had age-related neurodegenerative and/or cerebrovascular disease microscopic findings (Tables 3 and 4). For 5 BSHRI subjects, findings were limited to “primary age-related tauopathy” (PART) while the remainder of the BSHRI cases and all of the Mayo Clinic cases met clinicopathological diagnostic criteria for one or more defined entities, including Alzheimer’s dementia (AD, 10 cases), chronic cerebral white matter rarefaction or leukoencephalopathy (CWMR, 2 cases), dementia with Lewy bodies, diffuse Lewy body disease or transitional Lewy body disease (DLB, DLBD, TLDB, respectively, 5 total cases), limbic-predominant age-related TDP-43 encephalopathy or frontotemporolobar degeneration with TDP-43 proteinopathy (LATE and FTLD-TDP, respectively, 5 total cases), progressive supranuclear palsy (PSP, 5 cases), vascular dementia or atherosclerotic cerebrovascular disease (VaD, ASCVD, 4 cases), aging-related tau astrogliopathy or thorn-shaped astrocytes (ARTAG, TSA, respectively, 3 total cases), pallido-nigro-luysian atrophy (PNLA, 2 cases) and hippocampal sclerosis (HS, 1 case).

Only two subjects had microscopic findings that were definitely attributable to COVID-19 disease. One Mayo Clinic case had encephalitis, with perivascular mononuclear cell cuffing, acute hemorrhages, fibrinoid vascular necrosis and transtentorial temporal lobe uncal herniation (Case M10, Figures 1 - 3). One BSHRI subject (Case B3) had a large acute middle cerebral artery territory ischemic and hemorrhagic infarction (Figures 4 and 5) that was clinically documented to have occurred several days after the clinical onset of his COVID-19 disease, and was accompanied by bilateral lower extremity arterial thromboses. Other subjects had findings that may have been directly related to COVID-19 disease but that are also common in autopsies of elderly persons (Figures 6 and 7). Of BHSRI cases, B8 had acute microscopic infarcts, and cases B9 and B10 had acute microscopic hemorrhages. Case B6 had two microglial nodules in the posterior medulla and several BSHRI cases had widely-separated, multifocal sparse to moderate perivascular cuffing with mononuclear inflammatory cells, but without further infiltration of the neuropil. Seven BSHRI cases and four Mayo Clinic cases had multifocal white matter sites with APP-immunoreactive axonal swellings (Figure 3C-E), consistent with subacute or acute axonal damage, similarly to other recently reported cases of COVID-19 ^53,56^. Several BSHRI cases had mineralization of blood vessels within the globus pallidus, a common incidental autopsy finding in elderly brains, but in one (Case B8) this was extensive and also involved the dentate gyrus, cerebellar white matter and a laminar distribution of neurons in the occipital cortex (Figure 6E, F). Acute or subacute infarctions, ischemic changes or microhemorrhages were not present in any other subjects and otherwise there were no microscopic findings that might be unequivocally attributed to COVID-19 disease. Specifically, aside from the one Mayo Clinic case with encephalitis, no other case had convincing findings of viral infection, with no evidence of leptomeningitis or encephalitis, necrosis, perivascular or focal demyelination or viral intracellular inclusions.

**Figure 1.**
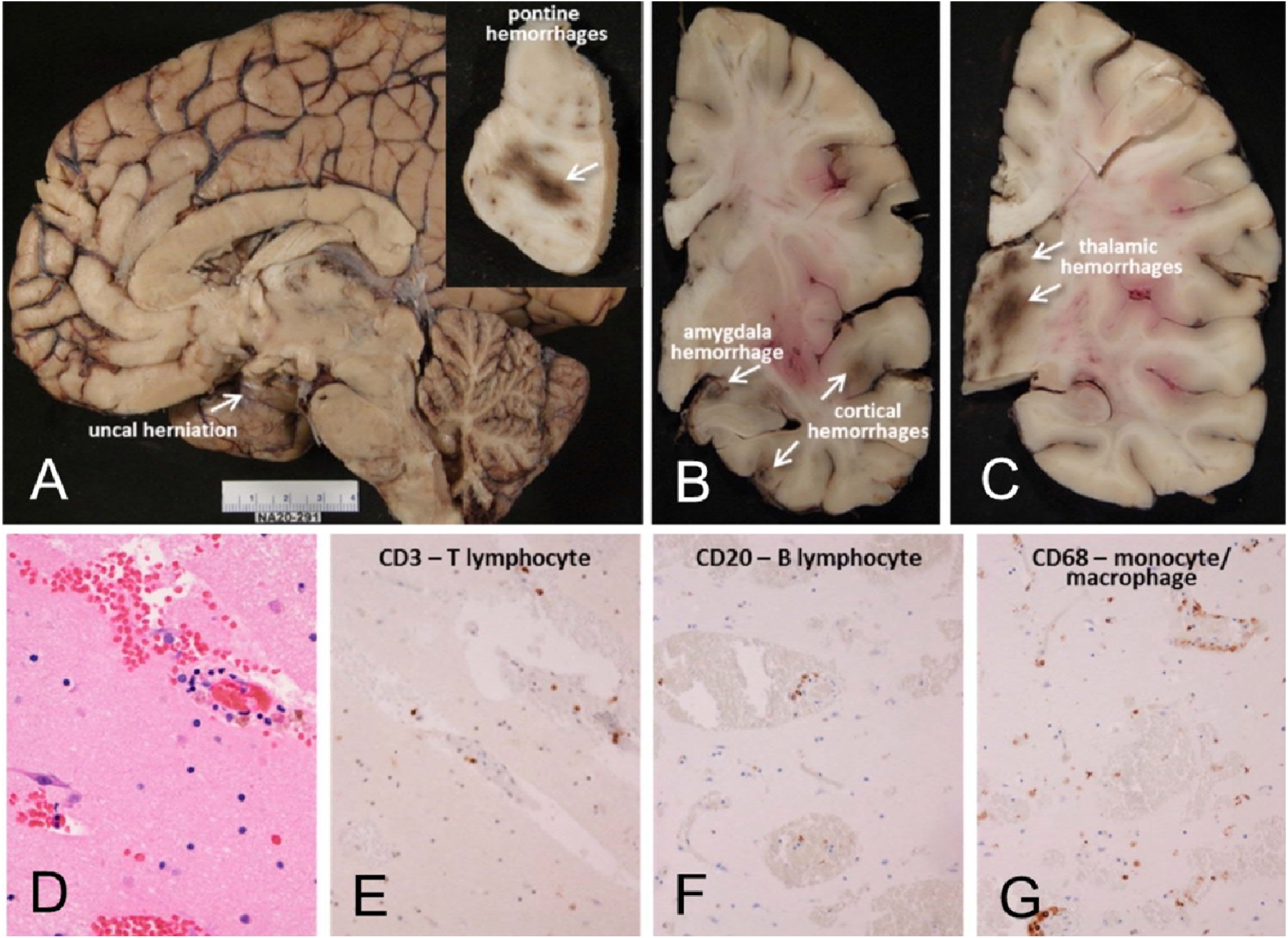
Mayo Clinic Case M10 (see Tables), a male in his 30s. Neuropathological gross examination was consistent with encephalitis-associated brain swelling causing transtentorial uncal herniation (A) and associated with acute hemorrhages (arrows A inset, and in B and C). Microscopic examination of semi-adjacent temporal lobe sections show microscopic hemorrhage (D; H & E stain) and neuropil infiltration with T- and B-lymphocytes and macrophages (E-G).

**Figure 2.**
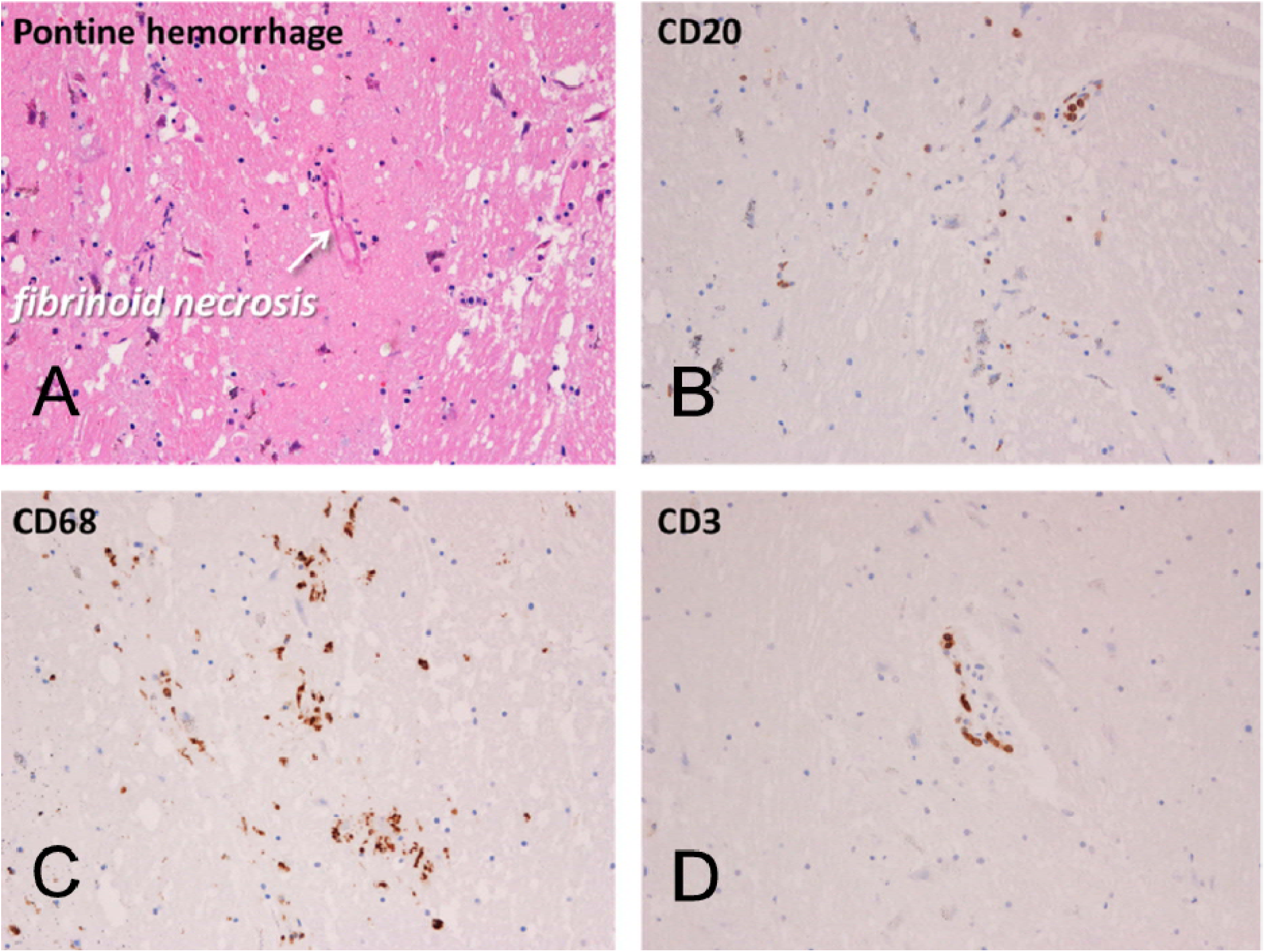
Mayo Clinic Case M10 (continued from Figure 1). Sections of pons adjacent to area of gross acute hemorrhage (Figure 1A inset). Acute pontine hemorrhage and fibrinoid necrosis of a blood vessel in a section stained with H & E (A). Semi-adjacent sections are immunohistochemically stained for B-lymphocytes (B, CD20), macrophages (C, CD68) and T-lymphocytes (D, CD3).

**Figure 3.**
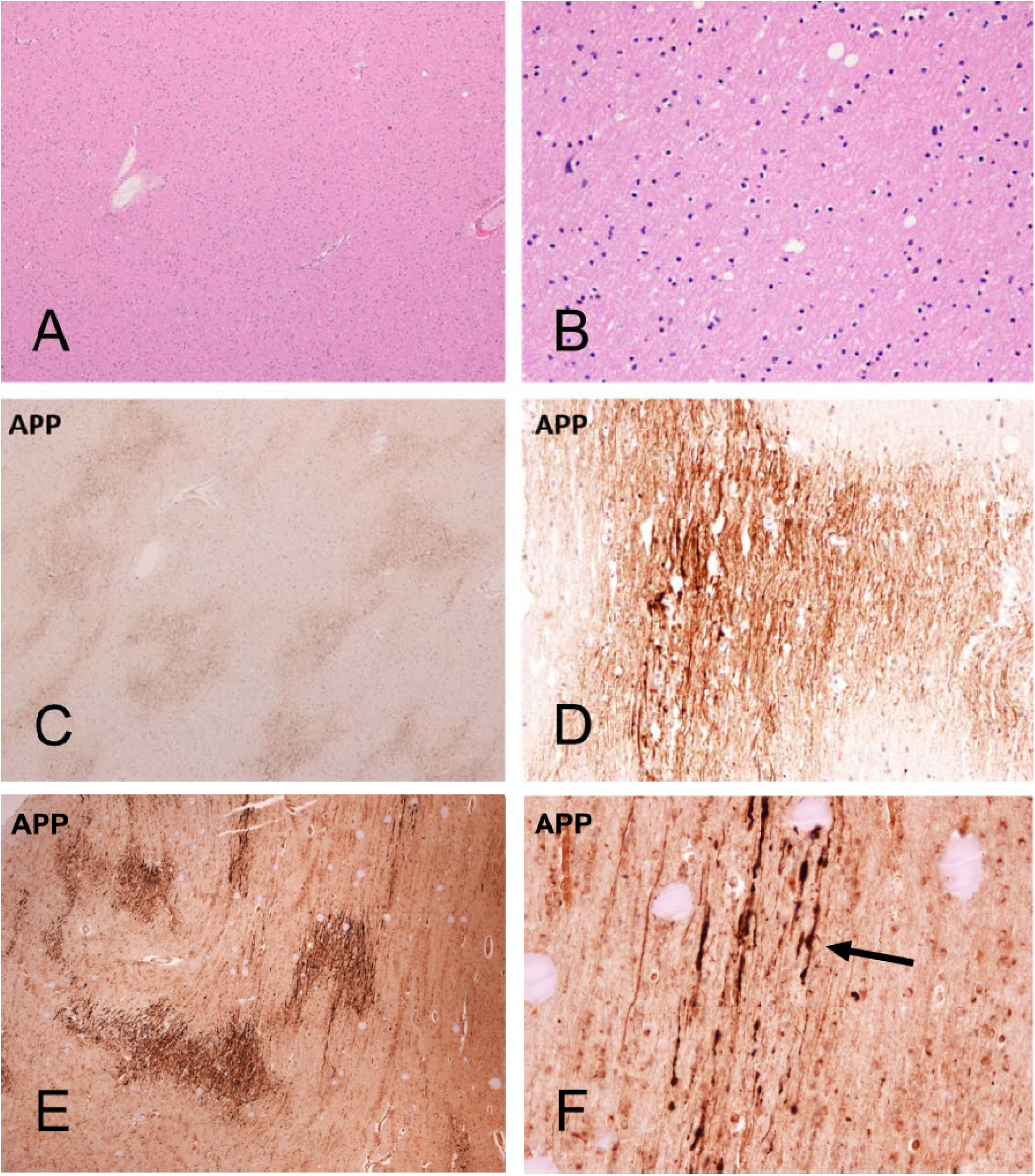
Cerebral white matter sections from Mayo Clinic Case M10 (continued from Figures 1 and 2, A-D) and BSHRI Case B5 (E, F), a male in his 90s. Sections of cerebral white matter from Case M10 are unremarkable on H & E stains, at low (A) and medium (B) magnifications while semi-adjacent sections stained with an immunohistochemical method for β-amyloid precursor protein (APP) show patchy staining at lo magnification and intense staining of axons at higher magnification (C and D, respectively). Lower (E) nd higher (F) magnifications of APP immunostaining in Case B5b show a patchy quality at lower magnification, with axonal staining visible at higher magnifications. Axons frequently have periodic axonal swellings (arrow in F shows example).

**Figure 4.**
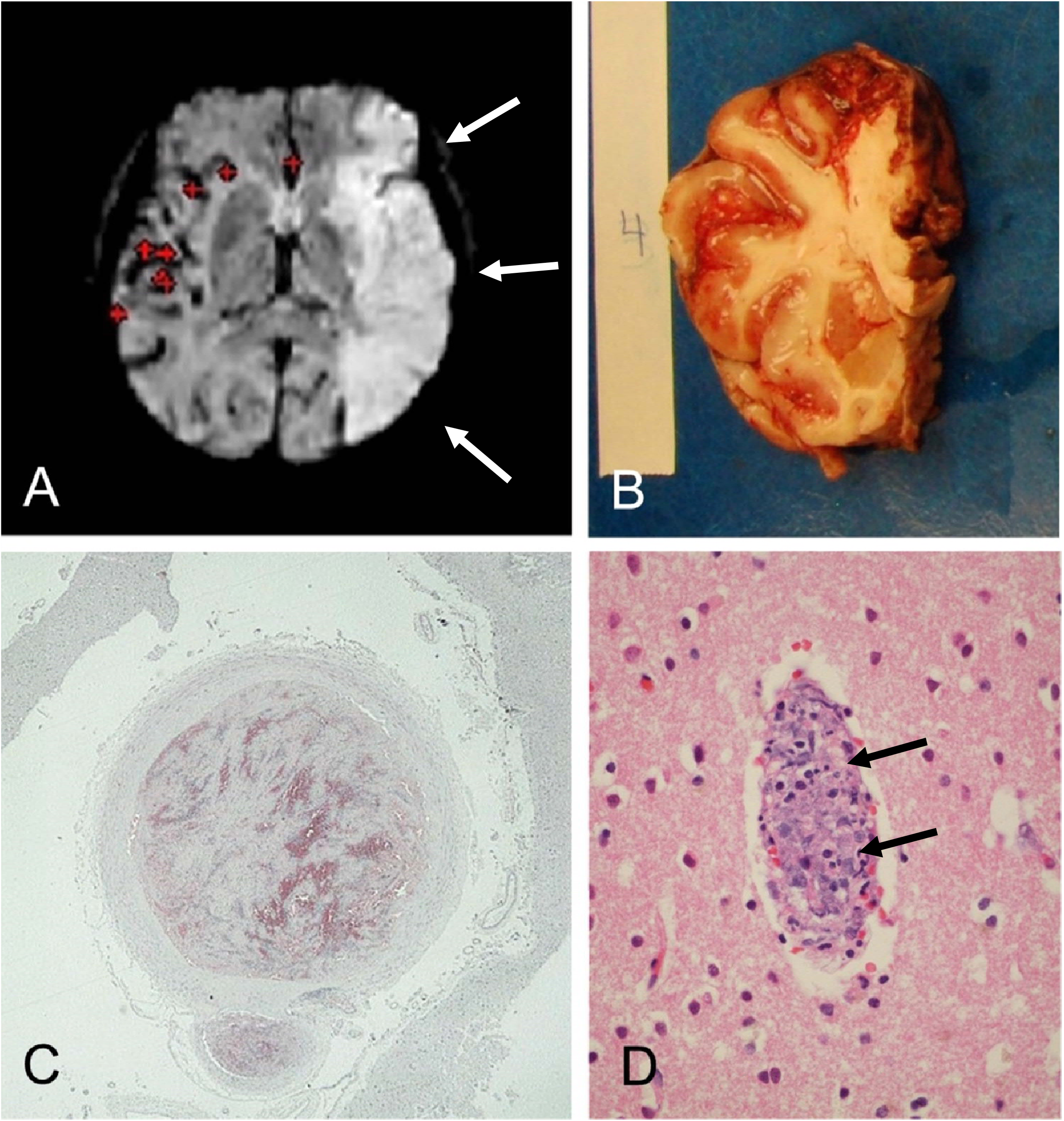
Banner Sun Health Research Institute (BSHRI) Case B3, a male in his 70s. Clinical findings indicated a massive acute left middle cerebral artery territory ischemic infarct, as shown in the MRI image (A, arrows). Gross examination of the brain at autopsy showed widespread hemorrhagic areas, especially in cerebral cortex (B). The left middle cerebral artery within the Sylvian fissure was completely occluded by firm thrombus, confirmed on microscopic examination (C). There were multiple thrombi within parenchymal arterioles (D, arrows). Sections were stained with H & E, on 6 μm paraffin sections

**Figure 5.**
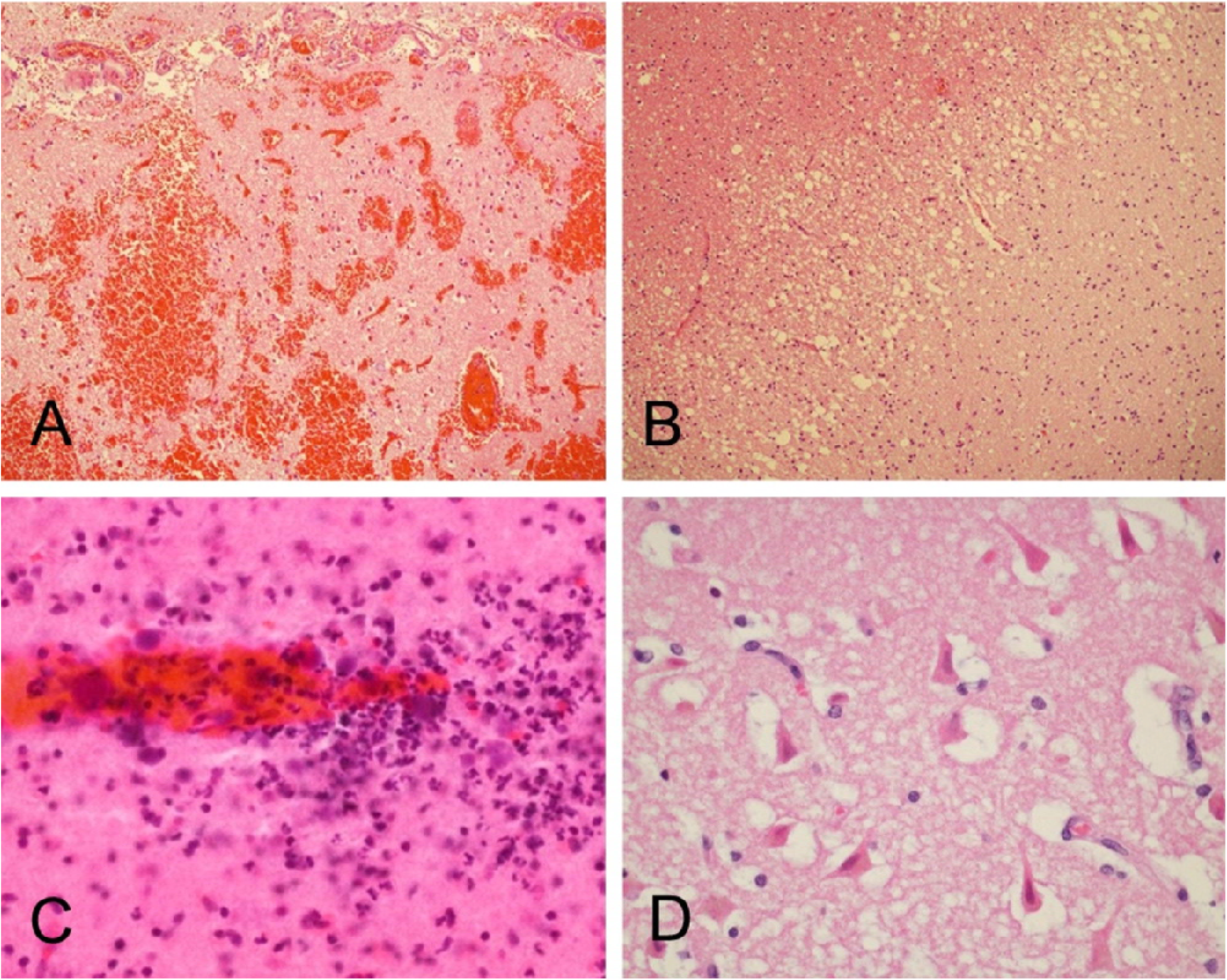
Case B3 (continued from Figure 4). Microscopic examination confirmed the presence of widespread acute cortical hemorrhages (A). Other areas within the left middle cerebral artery distribution showed acute ischemic infarction, with microvacuolation of the neuropil and loss of normal tissue eosinophilia (B), perivascular neuropil infiltration by polymorphonuclear leukocytes (C), and acute hypoxic-ischemic changes, including perikaryal cytoplasmic eosinophilia and nuclear pyknosis of cortical pyramidal neurons (D). Sections are all stained with H & E, on 6 μm paraffin sections (A, B, D) or 80 μm thick sections (C)

**Figure 6.**
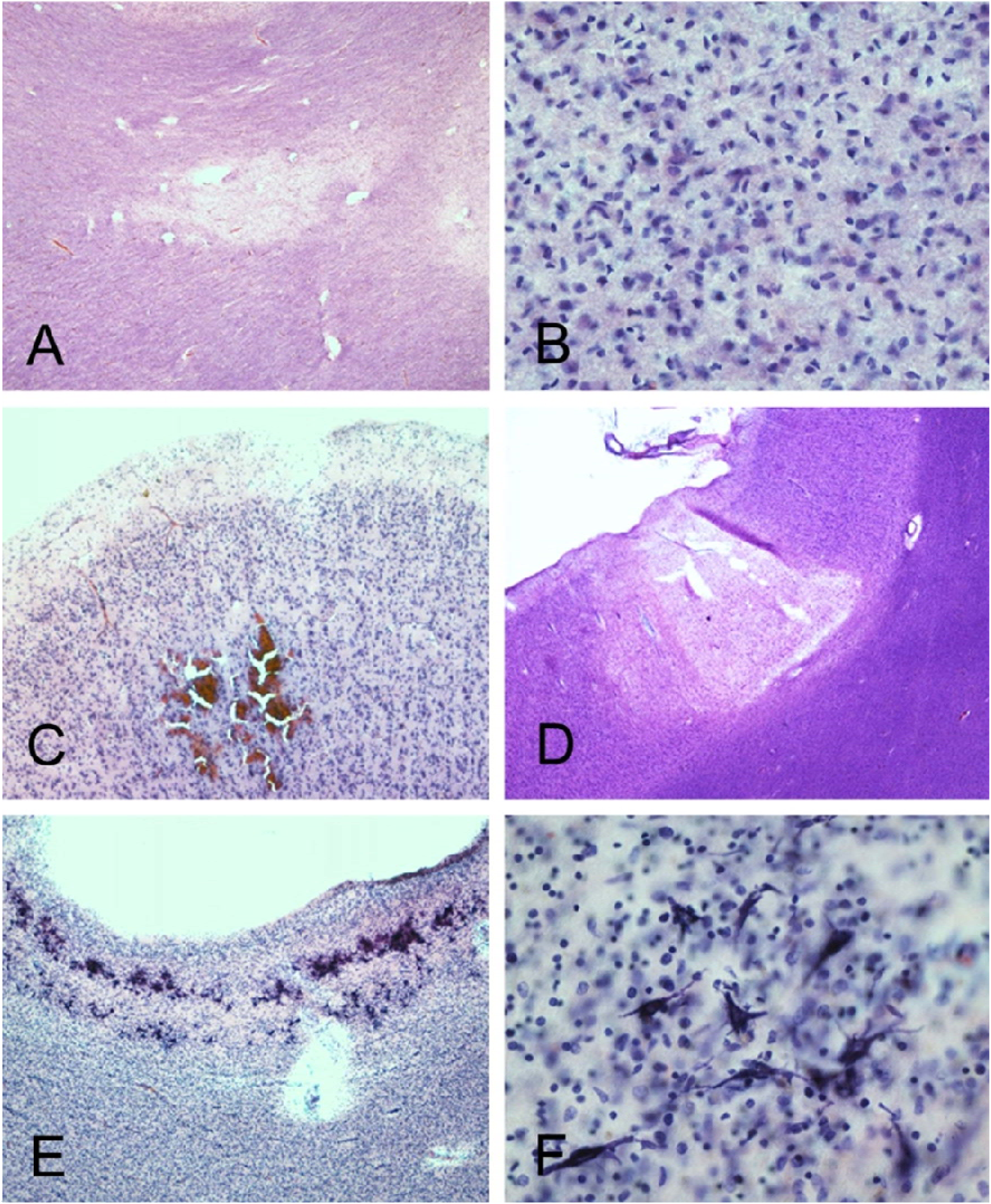
BSHRI Case B1 (A and B), a male in his 90s. Microscopic examination showed focal white matter rarefaction in the temporal lobe white matter (A) with increased numbers of microglial nuclei within area of rarefaction (B). BSRHI Case B9, a malein his 70s, showed acute microhemorrhages, seen here in the cortex of the superior frontal gyrus (C). BSHRI Case B8 (D-F), a male in his 80s, had an acute microscopic infarct in cortex of the middle frontal gyrus (D) and laminar mineralization of pyramidal neurons (E, F) in lateral occipital association cortex. All images are from H & E-stained 80 μm thick sections.

**Figure 7.**
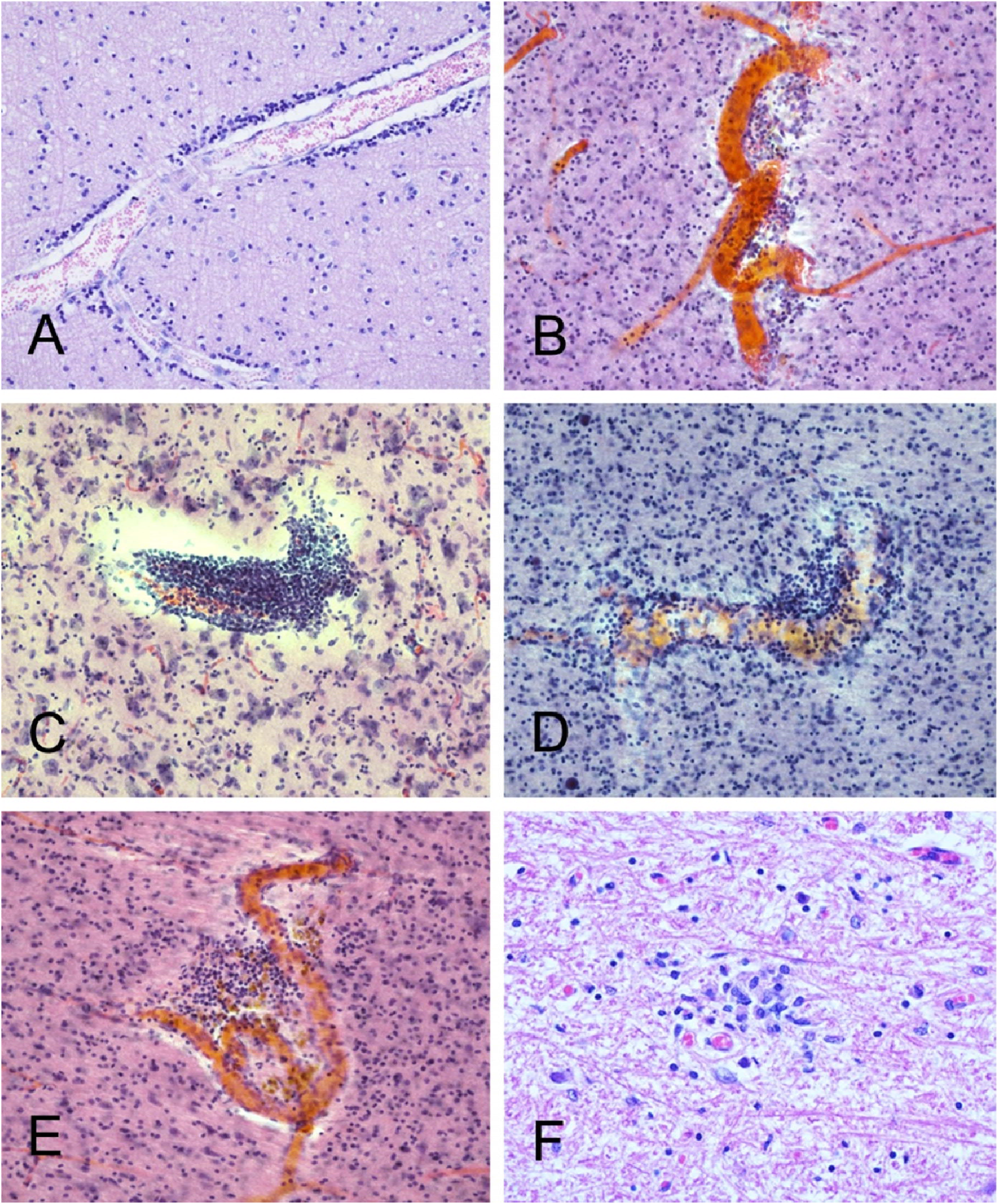
Several cases had occasional perivascular mononuclear cell aggregates, in both gray and white matter. Shown are examples from BSHRI Case B4, a male in his 80s(A), Case B2, a female in her 70s (B), Case B6, a male in his 70s (C), Case B8, a male in his 80s(D), and Case B9, a male in his 70s (E). Also shown is a microglial nodule in the posterior medulla in the region of the nucleus gracilis, in BSHRI Case B6 (F). Images are from H & E-stained 80 μm thick sections (B-E) or 6 μm paraffin sections (A, F).

### SARS-CoV-2 RT-PCR Findings

Eight of nine of the BSHRI COVID-19 disease cases with whole-body autopsy showed above-threshold amplification of the SCV2 E gene in their lung samples (Table 5) but only two of the ten BSHRI cases, B9 and B10, amplified beyond threshold in any of the 16 brain regions sampled (Table 5 and Supplemental Table 5). For Case B9, amplification was seen only in the dorsal medulla and the olfactory bulb. For case B10, amplification was seen in the amygdala as well as the frontal and temporal neocortex. Interpolating from a synthetic SCV2 standard concentration curve (Figure 8) we estimated that the heaviest sample viral load was approximately 189 copies of the E gene per µl, in the olfactory bulb of Case B9 while the lowest load was 0.7 copies per µl, in the entorhinal area of case M10. Brain loads were generally lower than those in the lungs, where the highest viral load was estimated at 300,000 copies per µl (Figure 8, Case B10). Case B9 had similar viral loads in lung and olfactory bulb. Case B3, the only BSHRI case with neuropathological findings that could be unequivocally attributed to COVID-19 disease (acute middle cerebral artery territory acute infarction and hemorrhages), did not reach threshold amplification in any of the 16 tested brain regions.

**Table 5.** Case data for SCV2 RNA detection in the brain and lung. Only samples considered positive for threshold amplification in the presence of adequate housekeeping gene amplification are shown (shaded cells). See Supplementary Table 5 for full results, including all negative results and results for CSF and blood serum. Case B1 and Cases M1-11 did not have lung samples available. Case B8 was PCR-negative in lung (shown only in Supplementary Table 5). CT= cycle threshold; E gene = envelope (*E*) gene; RNASE P = ribonucleus P gene; AMYG = amygdala; DM = dorsal medulla; ENT = entorhinal area; LEPTOS = leptomeninges; FC = frontal neocortex; TC = temporal neocortex; OB = olfactory bulb.

| Case | Sample | E Gene Ct | RNASE P Ct |
| --- | --- | --- | --- |
| B2 | Lung | 35 | 24 |
| B3 | Lung | 30 | 24 |
| B4 | Lung | 25 | 23 |
| B5 | Lung | 28 | 24 |
| B6 | Lung | 35 | 24 |
| B7 | Lung | 31 | 23 |
| B9 | DM | 40 | 24 |
| B9 | Lung | 28 | 24 |
| B9 | OB | 28 | 23 |
| B10 | AMYG | 35 | 23 |
| B10 | FC | 34 | 23 |
| B10 | Lung | 18 | 24 |
| B10 | TC | 30 | 23 |
| M10 | ENT | 36 | 24 |
| M10 | LEPTOS | 34 | 24 |
| M11 | OB | 29 | 19 |

**Figure 8.**
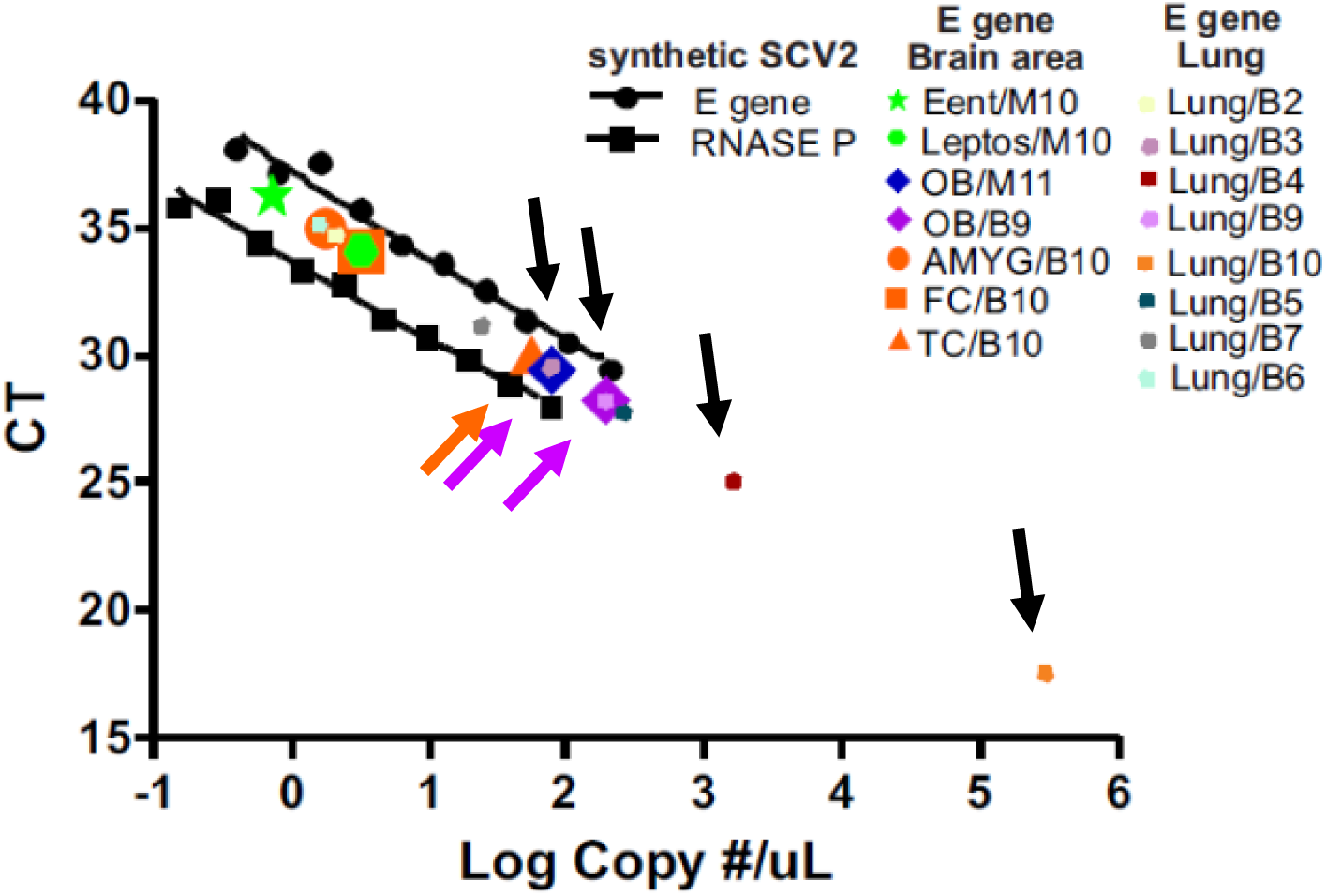
Standard curve for obtaining estimated SARS-CoV-2 sample copy numbers, constructed with serial dilutions of commercially-obtained synthetic SCV2 RNA. Ct values for patient samples that were positive for threshold amplification (colored symbols, see legend; some symbols are superimposed on others on graph) of sample SCV2 RNA are plotted on the same graph. The highest copy numbers were obtained from lung (black arrows show lung samples with higher Ct values), olfactory bulb (OB, lavender arrows show both of the positive OB samples) and temporal cortex (TC, orange arrow shows the only positive TC sample). Ent = entorhinal area; Leptos = leptomeninges; AMYG = amygdala; FC = frontal cortex.

Of the ten Mayo Clinic COVID-19 cases, again only two (Cases M10 and M11) showed above-threshold amplification of the SCV2 E gene in any brain region. For case M10, this was seen in the entorhinal and leptomeningeal samples, while for case M11, this was only in the olfactory bulb. Notably, case M10 was the only case with both a positive RT-PCR signal as well as neuropathological findings (encephalitis) that could be unequivocally attributed to COVID-19 disease.

The brain region with the highest viral load by cycle number, for both the BSHRI and Mayo Clinic cases, was the olfactory bulb.

None of the postmortem cardiac blood serum and cerebrospinal fluid samples, assayed in 8 and 9 BSHRI cases, respectively, had E gene amplification meeting the Ct 40 threshold.

Housekeeping gene (RNAase P and actin) amplification was adequate for all samples and serial amplified dilutions are plotted in Figure 8.

Threshold E gene amplification was not achieved in any of the lung, brain or biofluid samples from the 4 BSHRI non-COVID-19 control cases (Cases B11-B14).

## DISCUSSION

The findings of this study are generally in agreement with previously reported autopsy examinations of the brains of COVID-19 disease subjects, in that serious complications like encephalitis and large acute infarctions were present in relatively few subjects. For both of these features, 1 of 20, or 5% of subjects in the current study, were affected. Acute macroscopic or microscopic hemorrhages were present in four subjects (B3, B9, B10, M10) for a prevalence of 20% ^38–44,44–57^. Aside from the single Mayo Clinic encephalitis case (M10), and one BSHRI case (B6) with two microglial nodules, there was no clear evidence of the classical neuropathology of viral CNS infections ^58,59^, with no lymphocytic leptomeningitis or encephalitis, microglial nodules, pronounced or frequent perivascular lymphocytic cuffing, focal demyelination or viral inclusions.

Also in agreement with prior studies ^36–39,45,47–50,60–67^ is the relatively low rate of SCV2 RNA sequence presence in brain tissue, in 4 of 20 subjects (20%) in the current study, as compared with a mean of 22% (SD 32.7) across the total subjects included in 16 previously published studies, all with less-extensive brain sampling. The rate of SCV2 brain invasion in the current study has additional confidence, relative to previous reports, due to the large number of brain regions assayed, 16 brain regions per subject, in 20 subjects, for a total of 320 separate samples. Supporting a conclusion that the positive brain samples represent true brain invasion are the negative blood serum and CSF RT-PCR results for all 9 of the BSHRI cases that had whole-body autopsy, including the two with positive brain results; this makes it unlikely that the positive brain results were only coincidental to the presence of blood or CSF in the samples. Of the two cases with unequivocal and dramatic neuropathological findings, only one, the Mayo Clinic encephalitis case, was also positive for SCV2 RNA, and was so in a brain region affected by encephalitis (entorhinal area). The only case with microglial nodules, BSHRI case B6, was negative for SCV2 RNA.

These results confirm the growing consensus that most of the reported COVID-19 associated neurological signs and symptoms may be due, not to direct viral brain invasion, but to systemic reactions such as coagulopathy, sepsis, autoimmune mechanisms or multiorgan failure ^36^. The BSHRI case with acute cerebral ischemic and hemorrhagic infarction was likely an example of a systemic COVID-19-associated coagulopathy affecting the brain, as this subject also had extensive arterial thromboses of the lower extremities. The rare occurrence of virus in the brain contrasts with the commonly observed neurological consequences of COVID-19, reinforcing the concept that mitigating host response dysregulations (in coagulation, inflammation, autoimmunity, and more) may be key to limiting pathophysiology due to COVID-19 and other severe respiratory infections.

An important new contribution of the present study is the neuroanatomically-detailed SCV2 brain mapping, which may provide insights into the routes by which the virus enters the brain. We had hypothesized, as have others ^1,4,9,49,64,71,72,76–78,83^ that SCV2 RNA would most often be found in those brain regions implicated as environmental entry zones and areas with neuroanatomically-close connections to these, and therefore we chose to assay the olfactory bulb, adjacent to the nasopharyngeal mucosa, and its closely-connected regions including amygdala, hippocampus and entorhinal area, as well as the trigeminal and vagus nerve entry zones in the pontine tegmentum and dorsal medulla. We further hypothesized that other brain regions more distant would be less often involved and/or would have lower SCV2 viral loads and thus we tested for SCV2 in representatives of such areas, including cerebellar cortex, thalamus and lentiform nucleus. This hypothesis was supported to some extent as olfactory bulb was the only region with a positive PCR signal in more than one subject, and had the strongest PCR signals of any brain area, while of remaining positive brain regions, amygdala and entorhinal areas were each positive in one subject. Of cerebral neocortical areas, frontal and temporal lobes were each positive in one subject but there was no positive result for more distant regions including the primary visual area and cerebellar cortex. The dorsal medulla was positive in only a single case while the pontine tegmentum was not positive in any case. Tempering the confidence of any conclusions that might be reached, however, is the small number of positive results for each brain region, the focality of sampling within each region, the presence of brainstem or olfactory bulb positivity in only 1 and 2 of the 4 cases with positive brain signals, respectively, and the lack of any specific histopathological findings for viral CNS infection (except for case M10) in these brain regions. Information on olfactory function changes as part of our subjects’ COVID-19 illness is not available.

We had hypothesized that SCV2 brain invasion, as with some murine coronaviruses ^24,25,103^ might occur through the bloodstream, and so we tested sites that might be reflective of this, including the choroid plexus and leptomeninges, which are predominantly composed of blood vessels, as well as the hypothalamus and dorsal medulla, which contain regions (median eminence and area postrema) that lack a blood-brain-barrier. Single cases were positive in the leptomeninges and dorsal medulla, providing limited but non-conclusive support for a hematogenous entry.

Prior published studies have reported evidence that SCV2 brain invasion might occur through the olfactory bulb, vagus nerve or trigeminal nerve. A Basel-based group reported positive SCV2 PCR signals in 4 of 7 cases in olfactory bulb but no positives in brainstem regions^36^. A Hamburg group had positive signals in 4 of 8 cases in the medulla^61^. A Berlin group reported 3/31 cases positive in olfactory bulb, 1 of 7 in olfactory tubercle, 6/31 in medulla and 3/22 in trigeminal ganglion^49^. A Boston group had 3 of 16 cases positive in medulla and the same fraction in olfactory bulbs or tracts (in 5 different subjects)^67^.

Importantly, the CSF and blood serum were negative in all cases with samples available, making it unlikely that these would be useful for clinical SCV2 diagnosis. This is again in general agreement with several prior reports ^104–107^. The low rate of SCV2 infectivity in blood and circulating blood cells does not favor the existence of a substantial hematogenous pathway to the brain^108–111^.

Because of concerns that SCV2 might affect the substantia nigra (SN) and result in post-encephalitic parkinsonism ^112^, we assayed the SN but it was negative in all 20 cases. Whether CNS coronavirus persistence could lead to delayed clinical disease, either by viral reactivation or immune-mediated mechanisms, is unknown, but different strains of MHV, a coronavirus, can persist for prolonged periods in mice without any clinical expression or with chronic demyelination ^70,113–117^. If the 20% rate of SCV2 brain invasion we have documented here is a valid estimate for all those that have been infected, then with 20 million US COVID-19 cases there could be 4 million with the potential for long-term viral CNS persistence. This is likely to be an overestimate since the COVID-19 cases in this study were all severely affected while the majority of US cases have been asymptomatic or only mildly affected. Although chronic neurological sequelae of human coronavirus infections have not been unequivocally identified, this possibility should not be ignored. Coronaviruses have been suggested as a cause of human multiple sclerosis ^21^. Two human coronaviruses, strains 229E and OC43, have been demonstrated by RT-PCR, Northern hybridization and in-situ hybridization in 44% and 23%, respectively, of 90 human brains obtained from brain banks, indicating the potential for long-lasting CNS persistence after infection ^21^.

Many of the reported neuropathological findings in COVID-19 autopsies are of uncertain specificity, as they are also common in unselected autopsy series. Postmortem studies to date have rarely had a non-COVID-19 control group. While less often meeting criteria for frank infarction, several groups have reported the frequent presence in COVID-19 cases of acute hypoxic-ischemic changes ^48,53,55–57,67,118^ and it has been suggested that detection of swollen axons with immunohistochemical stains for APP might be more sensitive to such alterations or even be a “signature” change of hypoxic leukoencephalopathy specific to COVID-19 brain disease ^53,56^. The APP stain detected swollen white matter axons in 7 of 10 BSHRI cases and 5 of 10 Mayo Clinic cases. These APP-positive features, however, have been reported in association with a variety of conditions, including human cases and/or animal models of ischemia ^119,120^, traumatic brain injury ^121–123^, Binswanger’s disease or vascular dementia ^124,125^, bacterial meningitis ^126^ and drug abuse ^127^.

In the current study, sparse or moderate perivascular mononuclear chronic inflammatory cell aggregates were seen in almost all of the BSRHI cases but only in a single Mayo Clinic case, perhaps due to the larger brain volume sampled by the large-format 80 μm thick sections and hence larger brain sample volume employed by BSHRI. Such perivascular “cuffing” is a typical feature of viral brain infections but is also seen as a common incidental finding at autopsy. In no case except the single Mayo Clinic encephalitis case was there more extensive infiltration of the perivascular neuropil by these mononuclear cells and only in this Mayo Clinic case was there focal fibrinoid vascular necrosis. We did not find “endothelitis” to be a common feature, as described in one report^128^ and would find this difficult to distinguish from incidental postmortem capillary distention with polymorphonuclear leukocytes.

Calcification or mineralization of intraparenchymal blood vessels was prominent in one BSHRI case. This is common in elderly brains, and, as in the current case, most often involves the globus pallidus, dentate gyrus and dentate nucleus/deep cerebellar white matter. Frequently this is asymptomatic and is usually considered a chronic idiopathic change with many possible causes including viral infection ^129^ but case reports also exist of globus pallidus vascular mineralization occurring within the acute or subacute phase of infarction or sepsis.^130,131^ Similarly, neuronal mineralization or ferrugination, seen in the same BSHRI case with prominent vascular mineralization, is more often a late change but has been documented as early as 3 days after an infarction ^132^.

Careful comparison with neuropathological findings in subjects who did not have SCV2 infection will be critical for determining whether many common autopsy findings are substantially more likely in COVID-19 disease, and whether they may represent direct or indirect viral effects.

The relatively low brain prevalence of the SCV2 viral genome that we and most others have documented may possibly be due to our sampling at only selected brain locations and only at a single timepoint during COVID-19 illness. Viral detection rates are known to vary with stage and severity of illness and although all subjects died due to COVID-19, consistent with severe disease, we do not know the duration of illness for all. Although we sampled the brain much more extensively than what has been done in any previous study, the volume of brain sampled is still a very small fraction of total brain volume. The commonly-used RT-PCR methods may vary in sensitivity and specificity between centers, and may be insensitive to very low viral loads. False-negative SCV2 RT-PCR results may result from the presence of contaminating host RNase P DNA sequences that mask inadequate amounts of sample RNA^133^. Our RNA purification protocol included a gDNA-eliminating spin column step, reducing the likelihood of housekeeping gene host DNA amplification. Immunohistochemical or in-situ hybridization methods for SCV2 localization have been used by several groups^37,39,48,54,65,118,134^ but the significance or validity of such results are still unclear when RT-PCR assays of the same brain tissue regions are negative.

Similarly, the low rates of COVID-19-associated brain histopathology observed in our study and other studies might be increased if more sensitive or specific staining methods were used in all cases, including stains for microglial responses, as both human and experimental animal studies have shown that sepsis or its simulation, for example by peripheral lipopolysaccharide (LPS) injections, results in widespread CNS microglial activation that has been hypothesized as a cause of delirium^135^. A few groups have reported COVID-19 results with microglial stains ^61,67,118^.

Many of the subjects in this study had neurodegenerative diseases, due to the research focus of the participating academic centers. It is likely that these confer increased risk for COVID-19, beyond that due to age alone. The apolipoprotein E4 allele, a genetic risk factor for Alzheimer’s disease (AD), has been reported to be more common in subjects dying with severe COVID-19 disease ^136–138^, and non-COVID-19 associated pneumonia is reportedly more common in subjects with dementia or parkinsonism^139^. Several groups have reported greater prevalence and/or severity of COVID-19 disease in subjects with dementia or parkinsonism.^140–147^

Like other human coronaviruses, SARS-CoV-2 can inflict acute or subacute neuropathology in susceptible individuals. Much remains to be understood however, including what virus-host interactions influence CNS neuroinvasion and dissemination, and whether the virus persists or is cleared subsequent to acute illness. Further investigations will hopefully enable the development of improved surveillance, diagnostic and therapeutic strategies.

Biospecimens from the Banner Sun Health Research Institute Brain and Body Donation Program, including those presented in this report, are available to qualified researchers upon request at https://www.brainandbodydonationregistration.org/.

## Data Availability

Data will be available upon request.

https://www.brainandbodydonationprogram.org

## ACKNOWLEDGEMENTS AND FUNDING

This project was supported by a COVID-19 Supplement to a National Institute on Aging grant, (3P30AG019610-20S1), to the Arizona Alzheimer’s Disease Core Center, submitted in response to a Notice of Special Interest (NOSI) issued by the National Institute on Aging (NOT-AG-20-022), “to highlight the urgent need for research on Coronavirus Disease 2019…”.

## REFERENCES

1. Wu, Y, Xu, X, Chen, Z, et al. Nervous system involvement after infection with COVID-19 and other coronaviruses. Brain Behav Immun. 2020.

2. Gu, J and Korteweg, C. Pathology and pathogenesis of severe acute respiratory syndrome. Am J Pathol. 2007; 170:1136–1147.

3. Baig, AM, Khaleeq, A, Ali, U, et al. Evidence of the COVID-19 Virus Targeting the CNS: Tissue Distribution, Host-Virus Interaction, and Proposed Neurotropic Mechanisms. ACS Chem Neurosci. 2020; 11:995–998.

4. Lapina, C, Rodic, M, Pechanski, D, et al. The potential genetic network of human brain SARS-CoV-2 infection. BIORXIV. 2020; https://doi.org/10.1101/2020.04.06.027318.

5. McCray, PB, Jr., Pewe, L, Wohlford-Lenane, C, et al. Lethal infection of K18-hACE2 mice infected with severe acute respiratory syndrome coronavirus. J Virol. 2007; 81:813–821.

6. Ding, Y, He, L, Zhang, Q, et al. Organ distribution of severe acute respiratory syndrome (SARS) associated coronavirus (SARS-CoV) in SARS patients: implications for pathogenesis and virus transmission pathways. J Pathol. 2004; 203:622–630.

7. Gu, J, Gong, E, Zhang, B, et al. Multiple organ infection and the pathogenesis of SARS. J Exp Med. 2005; 202:415–424.

8. Xu, J, Zhong, S, Liu, J, et al. Detection of severe acute respiratory syndrome coronavirus in the brain: potential role of the chemokine mig in pathogenesis. Clin Infect Dis. 2005; 41:1089–1096.

9. Netland, J, Meyerholz, DK, Moore, S, et al. Severe acute respiratory syndrome coronavirus infection causes neuronal death in the absence of encephalitis in mice transgenic for human ACE2. J Virol. 2008; 82:7264–7275.

10. Li, K, Wohlford-Lenane, C, Perlman, S, et al. Middle East Respiratory Syndrome Coronavirus Causes Multiple Organ Damage and Lethal Disease in Mice Transgenic for Human Dipeptidyl Peptidase 4. J Infect Dis. 2016; 213:712–722.

11. Hung, EC, Chim, SS, Chan, PK, et al. Detection of SARS coronavirus RNA in the cerebrospinal fluid of a patient with severe acute respiratory syndrome. Clin Chem. 2003; 49:2108–2109.

12. Nilsson, A, Edner, N, Albert, J, et al. Fatal encephalitis associated with coronavirus OC43 in an immunocompromised child. Infect Dis (Lond). 2020;1–4.

13. Yeh, EA, Collins, A, Cohen, ME, et al. Detection of coronavirus in the central nervous system of a child with acute disseminated encephalomyelitis. Pediatrics. 2004; 113:e73–e76.

14. Morfopoulou, S, Brown, JR, Davies, EG, et al. Human Coronavirus OC43 Associated with Fatal Encephalitis. N Engl J Med. 2016; 375:497–498.

15. Murray, RS, Cai, GY, Hoel, K, et al. Coronavirus infects and causes demyelination in primate central nervous system. Virology. 1992; 188:274–284.

16. Wege, H, Winter, J, Korner, H, et al. Coronavirus induced demyelinating encephalomyelitis in rats: immunopathological aspects of viral persistency. Adv Exp Med Biol. 1990; 276:637–645.

17. Jacomy, H, St-Jean, JR, Brison, E, et al. Mutations in the spike glycoprotein of human coronavirus OC43 modulate disease in BALB/c mice from encephalitis to flaccid paralysis and demyelination. J Neurovirol. 2010; 16:279–293.

18. Bergmann, CC, Lane, TE, and Stohlman, SA. Coronavirus infection of the central nervous system: host-virus stand-off. Nat Rev Microbiol. 2006; 4:121–132.

19. Wang, FI, Stohlman, SA, and Fleming, JO. Demyelination induced by murine hepatitis virus JHM strain (MHV-4) is immunologically mediated. J Neuroimmunol. 1990; 30:31–41.

20. Trandem, K, Jin, Q, Weiss, KA, et al. Virally expressed interleukin-10 ameliorates acute encephalomyelitis and chronic demyelination in coronavirus-infected mice. J Virol. 2011; 85:6822–6831.

21. Arbour, N, Day, R, Newcombe, J, et al. Neuroinvasion by human respiratory coronaviruses. J Virol. 2000; 74:8913–8921.

22. Wege, H. Immunopathological aspects of coronavirus infections. Springer Semin Immunopathol. 1995; 17:133–148.

23. Houtman, JJ and Fleming, JO. Pathogenesis of mouse hepatitis virus-induced demyelination. J Neurovirol. 1996; 2:361–376.

24. Cabirac, GF, Soike, KF, Zhang, JY, et al. Entry of coronavirus into primate CNS following peripheral infection. Microb Pathog. 1994; 16:349–357.

25. Cabirac, GF, Murray, RS, McLaughlin, LB, et al. In vitro interaction of coronaviruses with primate and human brain microvascular endothelial cells. Adv Exp Med Biol. 1995; 380:79–88.

26. Murray, RS, Cai, GY, Soike, KF, et al. Further observations on coronavirus infection of primate CNS. J Neurovirol. 1997; 3:71–75.

27. Murray, RS, Cai, GY, Hoel, K, et al. Coronavirus infects and causes demyelination in primate central nervous system. Virology. 1992; 188:274–284.

28. Varatharaj, A, Thomas, N, Ellul, MA, et al. Neurological and neuropsychiatric complications of COVID- 19 in 153 patients: a UK-wide surveillance study. Lancet Psychiatry. 2020; 7:875–882.

29. Delamarre, L, Gollion, C, Grouteau, G, et al. COVID-19-associated acute necrotising encephalopathy successfully treated with steroids and polyvalent immunoglobulin with unusual IgG targeting the cerebral fibre network. J Neurol Neurosurg Psychiatry. 2020; 91:1004–1006.

30. Ghannam, M, Alshaer, Q, Al-Chalabi, M, et al. Neurological involvement of coronavirus disease 2019: a systematic review. J Neurol. 2020; 267:3135–3153.

31. Anand, PZLBNHDHGDMC-AAM. Neurologic Findings Among Inpatients with COVID-19 at a Safety-Net U.S. Hospital. Neurology Clin Practive. 2020; In press.

32. Kremer, S, Lersy, F, de, SJ, et al. Brain MRI Findings in Severe COVID-19: A Retrospective Observational Study. Radiology. 2020; 297:E242–E251.

33. Pons-Escoda, A, Naval-Baudin, P, Majos, C, et al. Neurologic Involvement in COVID-19: Cause or Coincidence? A Neuroimaging Perspective. AJNR Am J Neuroradiol. 2020; 41:1365–1369.

34. Romero-Sanchez, CM, Diaz-Maroto, I, Fernandez-Diaz, E, et al. Neurologic manifestations in hospitalized patients with COVID-19: The ALBACOVID registry. Neurology. 2020; 95:e1060–e1070.

35. Ellul, MA, Benjamin, L, Singh, B, et al. Neurological associations of COVID-19. Lancet Neurol. 2020; 19:767–783.

36. Deigendesch, N, Sironi, L, Kutza, M, et al. Correlates of critical illness-related encephalopathy predominate postmortem COVID-19 neuropathology. Acta Neuropathol. 2020; 140:583–586.

37. Lee, MH, Perl, DP, Nair, G, et al. Microvascular Injury in the Brains of Patients with COVID-19. N Engl J Med. 2020.

38. Jensen, MP, Le, QJ, Officer-Jones, L, et al. Neuropathological findings in two patients with fatal COVID-19. Neuropathol Appl Neurobiol. 2020.

39. Al-Sarraj, S, Troakes, C, Hanley, B, et al. Invited Review: The spectrum of neuropathology in COVID-19. Neuropathol Appl Neurobiol. 2020.

40. Pilotto, A, Odolini, S, Masciocchi, S, et al. Steroid-Responsive Encephalitis in Coronavirus Disease 2019. Ann Neurol. 2020; 88:423–427.

41. Poyiadji, N, Shahin, G, Noujaim, D, et al. COVID-19-associated Acute Hemorrhagic Necrotizing Encephalopathy: CT and MRI Features. Radiology. 2020;201187.

42. Moriguchi, T, Harii, N, Goto, J, et al. A first Case of Meningitis/Encephalitis associated with SARS-Coronavirus-2. Int J Infect Dis. 2020.

43. Mao, L, Jin, H, Wang, M, et al. Neurologic Manifestations of Hospitalized Patients With Coronavirus Disease 2019 in Wuhan, China. JAMA Neurol. 2020.

44. von Weyhern, CH, Kaufmann, I, Neff, F, et al. Early evidence of pronounced brain involvement in fatal COVID-19 outcomes. Lancet. 2020; 395:e109.

45. Freij, BJ, Gebara, BM, Tariq, R, et al. Fatal central nervous system co-infection with SARS-CoV-2 and tuberculosis in a healthy child. BMC Pediatr. 2020; 20:429.

46. Keller, E, Brandi, G, Winklhofer, S, et al. Large and Small Cerebral Vessel Involvement in Severe COVID-19: Detailed Clinical Workup of a Case Series. Stroke. 2020; 51:3719–3722.

47. Bihlmaier, K, Coras, R, Willam, C, et al. Disseminated Multifocal Intracerebral Bleeding Events in Three Coronavirus Disease 2019 Patients on Extracorporeal Membrane Oxygenation As Rescue Therapy. Crit Care Explor. 2020; 2:e0218.

48. Kantonen, J, Mahzabin, S, Mayranpaa, MI, et al. Neuropathologic features of four autopsied COVID-19 patients. Brain Pathol. 2020; 30:1012–1016.

49. Meinhardt, J, Radke, J, Dittmayer, C, et al. Olfactory transmucosal SARS-CoV-2 invasion as a port of central nervous system entry in individuals with COVID-19. Nat Neurosci. 2020.

50. Remmelink, M, De, MR, D’Haene, N, et al. Unspecific post-mortem findings despite multiorgan viral spread in COVID-19 patients. Crit Care. 2020; 24:495.

51. Bradley, BT, Maioli, H, Johnston, R, et al. Histopathology and ultrastructural findings of fatal COVID-19 infections in Washington State: a case series. Lancet. 2020; 396:320–332.

52. Borczuk, AC, Salvatore, SP, Seshan, SV, et al. COVID-19 pulmonary pathology: a multi-institutional autopsy cohort from Italy and New York City. Mod Pathol. 2020; 33:2156–2168.

53. Reichard, RR, Kashani, KB, Boire, NA, et al. Neuropathology of COVID-19: a spectrum of vascular and acute disseminated encephalomyelitis (ADEM)-like pathology. Acta Neuropathol. 2020; 140:1–6.

54. Al-Dalahmah, O, Thakur, KT, Nordvig, AS, et al. Neuronophagia and microglial nodules in a SARS-CoV-2 patient with cerebellar hemorrhage. Acta Neuropathol Commun. 2020; 8:147.

55. Conklin, J, Frosch, MP, Mukerji, S, et al. Cerebral Microvascular Injury in Severe COVID-19. medRxiv.2020.

56. Jaunmuktane, Z, Mahadeva, U, Green, A, et al. Microvascular injury and hypoxic damage: emerging neuropathological signatures in COVID-19. Acta Neuropathol. 2020; 140:397–400.

57. Song, E, Zhang, C, Israelow, B, et al. Neuroinvasion of SARS-CoV-2 in human and mouse brain. bioRxiv. 2020.

58. Ludlow, M, Kortekaas, J, Herden, C, et al. Neurotropic virus infections as the cause of immediate and delayed neuropathology. Acta Neuropathol. 2016; 131:159–184.

59. Seilhean, D. Infections of the central nervous system: Neuropathology. Rev Neurol (Paris). 2019; 175:431–435.

60. Deinhardt-Emmer, S, Wittschieber, D, Sanft, J, et al. Early postmortem mapping of SARS-CoV-2 RNA in patients with COVID-19 and correlation to tissue damage. bioRxiv. 2020;2020.

61. Matschke, J, Lutgehetmann, M, Hagel, C, et al. Neuropathology of patients with COVID-19 in Germany: a post-mortem case series. Lancet Neurol. 2020; 19:919–929.

62. Wichmann, D, Sperhake, JP, Lutgehetmann, M, et al. Autopsy Findings and Venous Thromboembolism in Patients With COVID-19: A Prospective Cohort Study. Ann Intern Med. 2020; 173:268–277.

63. Paniz-Mondolfi, A, Bryce, C, Grimes, Z, et al. Central nervous system involvement by severe acute respiratory syndrome coronavirus-2 (SARS-CoV-2). J Med Virol. 2020; 92:699–702.

64. Menter, T, Haslbauer, JD, Nienhold, R, et al. Postmortem examination of COVID-19 patients reveals diffuse alveolar damage with severe capillary congestion and variegated findings in lungs and other organs suggesting vascular dysfunction. Histopathology. 2020; 77:198–209.

65. Puelles, VG, Lutgehetmann, M, Lindenmeyer, MT, et al. Multiorgan and Renal Tropism of SARS-CoV- 2. N Engl J Med. 2020; 383:590–592.

66. Skok, K, Stelzl, E, Trauner, M, et al. Post-mortem viral dynamics and tropism in COVID-19 patients in correlation with organ damage. Virchows Arch. 2020.

67. Solomon, IH, Normandin, E, Bhattacharyya, S, et al. Neuropathological Features of COVID-19. N Engl J Med. 2020; 383:989–992.

68. Komaroff, AL, Pellett, PE, and Jacobson, S. Human Herpesviruses 6A and 6B in Brain Diseases: Association versus Causation. Clin Microbiol Rev. 2020; 34.

69. Abdullahi, AM, Sarmast, ST, and Singh, R. Molecular Biology and Epidemiology of Neurotropic Viruses. Cureus. 2020; 12:e9674.

70. Desforges, M, Le, CA, Dubeau, P, et al. Human Coronaviruses and Other Respiratory Viruses: Underestimated Opportunistic Pathogens of the Central Nervous System? Viruses. 2019; 12.

71. Desforges, M, Le, CA, Dubeau, P, et al. Human Coronaviruses and Other Respiratory Viruses: Underestimated Opportunistic Pathogens of the Central Nervous System? Viruses. 2019; 12.

72. Mori, I. Transolfactory neuroinvasion by viruses threatens the human brain. Acta Virol. 2015; 59:338–349.

73. Levinson, R, Elbaz, M, Ben-Ami, R, et al. Anosmia and dysgeusia appeared early in third of our patients with mild SARS-CoV-2 infection and were short-lived in most patients. MEDRXIV. 2020; https://doi.org/10.1101/2020.04.11.20055483.

74. Eliezer, M, Hautefort, C, Hamel, AL, et al. Sudden and Complete Olfactory Loss Function as a Possible Symptom of COVID-19. JAMA Otolaryngol Head Neck Surg. 2020.

75. Cantuti-Castelvetri, L, Ojha, R, Pedro, LD, et al. Neuropilin-1 facilitates SARS-CoV-2 cell entry and infectivity. Science. 2020; 370:856–860.

76. Kabbani, N and Olds, JL. Does COVID19 infect the brain? If so, smokers might be at a higher risk. Mol Pharmacol. 2020.

77. Butowt, R and Bilinska, K. SARS-CoV-2: Olfaction, Brain Infection, and the Urgent Need for Clinical Samples Allowing Earlier Virus Detection. ACS Chem Neurosci. 2020.

78. Hawrylycz, MJ, Lein, ES, Guillozet-Bongaarts, AL, et al. An anatomically comprehensive atlas of the adult human brain transcriptome. Nature. 2012; 489:391–399.

79. Doobay, MF, Talman, LS, Obr, TD, et al. Differential expression of neuronal ACE2 in transgenic mice with overexpression of the brain renin-angiotensin system. Am J Physiol Regul Integr Comp Physiol. 2007; 292:R373–R381.

80. Xia, H and Lazartigues, E. Angiotensin-converting enzyme 2 in the brain: properties and future directions. J Neurochem. 2008; 107:1482–1494.

81. Lin, Z, Chen, Y, Zhang, W, et al. RNA interference shows interactions between mouse brainstem angiotensin AT1 receptors and angiotensin-converting enzyme 2. Exp Physiol. 2008; 93:676–684.

82. Bohmwald, K, Galvez, NMS, Rios, M, et al. Neurologic Alterations Due to Respiratory Virus Infections. Front Cell Neurosci. 2018; 12:386.

83. Park, CH, Ishinaka, M, Takada, A, et al. The invasion routes of neurovirulent A/Hong Kong/483/97 (H5N1) influenza virus into the central nervous system after respiratory infection in mice. Arch Virol. 2002; 147:1425–1436.

84. Liu, JM, Tan, BH, Wu, S, et al. Evidence of central nervous system infection and neuroinvasive routes, as well as neurological involvement, in the lethality of SARS-CoV-2 infection. J Med Virol. 2020.

85. Dube, M, Le, CA, Wong, AHM, et al. Axonal Transport Enables Neuron-to-Neuron Propagation of Human Coronavirus OC43. J Virol. 2018; 92.

86. LeCoupanec A, Desforges, M, Meessen-Pinard, M, et al. Cleavage of a Neuroinvasive Human Respiratory Virus Spike Glycoprotein by Proprotein Convertases Modulates Neurovirulence and Virus Spread within the Central Nervous System. PLoS Pathog. 2015; 11:e1005261.

87. Beach, TG, Adler, CH, Sue, LI, et al. Arizona Study of Aging and Neurodegenerative Disorders and Brain and Body Donation Program. Neuropathology. 2015; 35:354–389.

88. Nelson, PT, Dickson, DW, Trojanowski, JQ, et al. Limbic-predominant age-related TDP-43 encephalopathy (LATE): consensus working group report. Brain. 2019; 142:1503–1527.

89. Dickson, DW, Rademakers, R, and Hutton, ML. Progressive supranuclear palsy: pathology and genetics. Brain Pathol. 2007; 17:74–82.

90. Dickson, DW, Ahmed, Z, Algom, AA, et al. Neuropathology of variants of progressive supranuclear palsy. Curr Opin Neurol. 2010; 23:394–400.

91. Crary, JF, Trojanowski, JQ, Schneider, JA, et al. Primary age-related tauopathy (PART): a common pathology associated with human aging. Acta Neuropathol. 2014; 128:755–766.

92. Kovacs, GG, Ferrer, I, Grinberg, LT, et al. Aging-related tau astrogliopathy (ARTAG): harmonized evaluation strategy. Acta Neuropathol. 2016; 131:87–102.

93. Roman, GC, Tatemichi, TK, Erkinjuntti, T, et al. Vascular dementia: diagnostic criteria for research studies. Report of the NINDS-AIREN International Workshop. Neurology. 1993; 43:250–260.

94. Mackenzie, IR, Neumann, M, Baborie, A, et al. A harmonized classification system for FTLD-TDP pathology. Acta Neuropathol. 2011; 122:111–113.

95. Gelb, DJ, Oliver, E, and Gilman, S. Diagnostic criteria for Parkinson disease. Arch Neurol. 1999; 56:33–39.

96. Dickson, DW, Braak, H, Duda, JE, et al. Neuropathological assessment of Parkinson’s disease: refining the diagnostic criteria. Lancet Neurol. 2009; 8:1150–1157.

97. Dickson, DW. Required techniques and useful molecular markers in the neuropathologic diagnosis of neurodegenerative diseases. Acta Neuropathol. 2005; 109:14–24.

98. Hyman, BT, Phelps, CH, Beach, TG, et al. National Institute on Aging-Alzheimer’s Association guidelines for the neuropathologic assessment of Alzheimer’s disease. Alzheimers Dement. 2012; 8:1–13.

99. Montine, TJ, Phelps, CH, Beach, TG, et al. National Institute on Aging-Alzheimer’s Association guidelines for the neuropathologic assessment of Alzheimer’s disease: a practical approach. Acta Neuropathol. 2012; 123:1–11.

100. McKeith, IG, Dickson, DW, Lowe, J, et al. Diagnosis and management of dementia with Lewy bodies: third report of the DLB Consortium. Neurology. 2005; 65:1863–1872.

101. Baba, Y, Ghetti, B, Baker, MC, et al. Hereditary diffuse leukoencephalopathy with spheroids: clinical, pathologic and genetic studies of a new kindred. Acta Neuropathol. 2006; 111:300–311.

102. Corman, VM, Landt, O, Kaiser, M, et al. Detection of 2019 novel coronavirus (2019-nCoV) by real-time RT-PCR. Euro Surveill. 2020; 25.

103. Bleau, C, Filliol, A, Samson, M, et al. Brain Invasion by Mouse Hepatitis Virus Depends on Impairment of Tight Junctions and Beta Interferon Production in Brain Microvascular Endothelial Cells. J Virol. 2015; 89:9896–9908.

104. Bellon, M, Schweblin, C, Lambeng, N, et al. Cerebrospinal fluid features in SARS-CoV-2 RT-PCR positive patients. Clin Infect Dis. 2020.

105. Lersy, F, Benotmane, I, Helms, J, et al. Cerebrospinal fluid features in COVID-19 patients with neurologic manifestations: correlation with brain MRI findings in 58 patients. J Infect Dis. 2020.

106. Placantonakis, DG, Aguero-Rosenfeld, M, Flaifel, A, et al. SARS-CoV-2 Is Not Detected in the Cerebrospinal Fluid of Encephalopathic COVID-19 Patients. Front Neurol. 2020; 11:587384.

107. Rosin, NL, Jaffer, A, Sinha, S, et al. SARS-CoV-2 infection of circulating immune cells is not responsible for virus dissemination in severe COVID-19 patients. bioRxiv. 2021;2021.

108. Politis, C, Papadaki, M, Politi, L, et al. Post-donation information and haemovigilance reporting for COVID-19 in Greece: Information supporting the absence of SARS-CoV-2 possible transmission through blood components. Transfus Clin Biol. 2020.

109. Rosin, NL, Jaffer, A, Sinha, S, et al. SARS-CoV-2 infection of circulating immune cells is not responsible for virus dissemination in severe COVID-19 patients. bioRxiv. 2021;2021.

110. Andersson, MI, Arancibia-Carcamo, CV, Auckland, K, et al. SARS-CoV-2 RNA detected in blood products from patients with COVID-19 is not associated with infectious virus. Wellcome Open Res. 2020; 5:181.

111. Kiely, P, Hoad, VC, Seed, CR, et al. Severe acute respiratory syndrome coronavirus-2: implications for blood safety and sufficiency. Vox Sang. 2020.

112. Brundin, P, Nath, A, and Beckham, JD. Is COVID-19 a Perfect Storm for Parkinson’s Disease? Trends Neurosci. 2020; 43:931–933.

113. Bender, SJ and Weiss, SR. Pathogenesis of murine coronavirus in the central nervous system. J Neuroimmune Pharmacol. 2010; 5:336–354.

114. Lane, TE and Hosking, MP. The pathogenesis of murine coronavirus infection of the central nervous system. Crit Rev Immunol. 2010; 30:119–130.

115. Elliott, R, Li, F, Dragomir, I, et al. Analysis of the host transcriptome from demyelinating spinal cord of murine coronavirus-infected mice. PLoS One. 2013; 8:e75346.

116. Talbot, PJ, Desforges, M, St-Jean, J, et al. Coronavirus neuropathogenesis: could SARS be the tip of the iceberg? BMC Proc. 2008; 2:S40.

117. Sorensen, O, Coulter-Mackie, MB, Puchalski, S, et al. In vivo and in vitro models of demyelinating disease. IX. Progression of JHM virus infection in the central nervous system of the rat during overt and asymptomatic phases. Virology. 1984; 137:347–357.

118. Schurink, B, Roos, E, Radonic, T, et al. Viral presence and immunopathology in patients with lethal COVID-19: a prospective autopsy cohort study. Lancet Microbe. 2020; 1:e290–e299.

119. Kalaria, RN, Bhatti, SU, Palatinsky, EA, et al. Accumulation of the beta amyloid precursor protein at sites of ischemic injury in rat brain. Neuroreport. 1993; 4:211–214.

120. MacKenzie, JM. Axonal Injury in Stroke: A Forensic Neuropathology Perspective. Am J Forensic Med Pathol. 2015; 36:172–175.

121. Hayashi, T, Ago, K, Nakamae, T, et al. Two different immunostaining patterns of beta-amyloid precursor protein (APP) may distinguish traumatic from nontraumatic axonal injury. Int J Legal Med. 2015; 129:1085–1090.

122. Ciallella, JR, Ikonomovic, MD, Paljug, WR, et al. Changes in expression of amyloid precursor protein and interleukin-1beta after experimental traumatic brain injury in rats. J Neurotrauma. 2002; 19:1555–1567.

123. Hortobagyi, T, Wise, S, Hunt, N, et al. Traumatic axonal damage in the brain can be detected using beta-APP immunohistochemistry within 35 min after head injury to human adults. Neuropathol Appl Neurobiol. 2007; 33:226–237.

124. Tomimoto, H, Lin, JX, Matsuo, A, et al. Different mechanisms of corpus callosum atrophy in Alzheimer’s disease and vascular dementia. J Neurol. 2004; 251:398–406.

125. Akiguchi, I, Tomimoto, H, Wakita, H, et al. Topographical and cytopathological lesion analysis of the white matter in Binswanger’s disease brains. Acta Neuropathol. 2004; 107:563–570.

126. Gerber, J, Seitz, RC, Bunkowski, S, et al. Evidence for frequent focal and diffuse acute axonal injury in human bacterial meningitis. Clin Neuropathol. 2009; 28:33–39.

127. Buttner, A, Rohrmoser, K, Mall, G, et al. Widespread axonal damage in the brain of drug abusers as evidenced by accumulation of beta-amyloid precursor protein (beta-APP): an immunohistochemical investigation. Addiction. 2006; 101:1339–1346.

128. Rhodes, RH, Love, GL, Da Silva Lameira, F, et al. Acute Endotheliitis (Type 3 Hypersensitivity Vasculitis) in Ten COVID-19 Autopsy Brains. medRxiv. 2021;2021.

129. Hawkins, CP, McLaughlin, JE, Kendall, BE, et al. Pathological findings correlated with MRI in HIV infection. Neuroradiology. 1993; 35:264–268.

130. Midroni, G and Willinsky, R. Rapid postanoxic calcification of the basal ganglia. Neurology. 1992; 42:2144–2146.

131. Mandal, AKJ, Patel, NB, and Missouris, CG. Sepsis Unmasking Fahr’s Disease. Am J Med Sci. 2020; 360:406–409.

132. Mena, H, Cadavid, D, and Rushing, EJ. Human cerebral infarct: a proposed histopathologic classification based on 137 cases. Acta Neuropathol. 2004; 108:524–530.

133. Dekker, RJ, Ensink, WA, van Leeuwen, S, et al. Overhauling a faulty control in the CDC-recommended SARS-CoV-2 RT-PCR test panel. bioRxiv. 2020;2020.

134. Radmanesh, A, Raz, E, Zan, E, et al. Brain Imaging Use and Findings in COVID-19: A Single Academic Center Experience in the Epicenter of Disease in the United States. AJNR Am J Neuroradiol. 2020; 41:1179–1183.

135. Hoogland, ICM, Westhoff, D, Engelen-Lee, JY, et al. Microglial Activation After Systemic Stimulation With Lipopolysaccharide and Escherichia coli. Front Cell Neurosci. 2018; 12:110.

136. Kuo, CL, Pilling, LC, Atkins, JL, et al. ApoE e4e4 Genotype and Mortality With COVID-19 in UK Biobank. J Gerontol A Biol Sci Med Sci. 2020; 75:1801–1803.

137. Kasparian, K, Graykowski, D, and Cudaback, E. Commentary: APOE e4 Genotype Predicts Severe COVID-19 in the UK Biobank Community Cohort. Front Immunol. 2020; 11:1939.

138. Nikogosov, DA, Shevlyakov, AD, and Baranova, AV. Comment on “ApoE e4e4 Genotype and Mortality With COVID-19 in UK Biobank” by Kuo et al. J Gerontol A Biol Sci Med Sci. 2020; 75:2233–2234.

139. Beach, TG. Increased Risk of Autopsy-Proven Pneumonia with Sex, Season and Neurodegenerative Disease. medRxiv. 2021.

140. Rutten, JJS, van Loon, AM, van, KJ, et al. Clinical Suspicion of COVID-19 in Nursing Home Residents: Symptoms and Mortality Risk Factors. J Am Med Dir Assoc. 2020; 21:1791–1797.

141. Espana, PP, Bilbao, A, Garcia-Gutierrez, S, et al. Predictors of mortality of COVID-19 in the general population and nursing homes. Intern Emerg Med. 2021.

142. Kim, SR, Nam, SH, and Kim, YR. Risk Factors on the Progression to Clinical Outcomes of COVID-19 Patients in South Korea: Using National Data. Int J Environ Res Public Health. 2020; 17.

143. Filardo, TD, Khan, MR, Krawczyk, N, et al. Comorbidity and clinical factors associated with COVID-19 critical illness and mortality at a large public hospital in New York City in the early phase of the pandemic (March-April 2020). PLoS One. 2020; 15:e0242760.

144. Hariyanto, TI, Putri, C, Arisa, J, et al. Dementia and outcomes from coronavirus disease 2019 (COVID- 19) pneumonia: A systematic review and meta-analysis. Arch Gerontol Geriatr. 2020; 93:104299.

145. Zhou, J, Liu, C, Sun, Y, et al. Cognitive disorders associated with hospitalization of COVID-19: Results from an observational cohort study. Brain Behav Immun. 2021; 91:383–392.

146. Atkins, JL, Masoli, JAH, Delgado, J, et al. Preexisting Comorbidities Predicting COVID-19 and Mortality in the UK Biobank Community Cohort. J Gerontol A Biol Sci Med Sci. 2020; 75:2224–2230.

147. Harrison, SL, Fazio-Eynullayeva, E, Lane, DA, et al. Comorbidities associated with mortality in 31,461 adults with COVID-19 in the United States: A federated electronic medical record analysis. PLoS Med. 2020; 17:e1003321.

