## Supplemental Table 1 for "Mapping of SARS-CoV-2 Brain Invasion and Histopathology in COVID-19 Disease"

**Supplementary Table 1.** Case data for SCV2 RNA detection in the brain, CSF and blood serum. Shaded cells are for assays considered positive. CT= cycle threshold; E gene= envelope (*E*) gene; RNASE P= ribonucleus P; A17= occipital primary visual neocortex; AMYG= amygdala; CBL= lateral cerebellar hemispheric cortex; CP= choroid plexus; CSF= cerebrospinal fluid; DM= dorsal medulla; ENT= entorhinal area; HIP= hippocampus; HYT= hypothalamus; LEPTOS= leptomeninges; LN= lentiform nucleus; FC= frontal neocortex; TC= temporal neocortex; OB= olfactory bulb; SN= substantia nigra; THAL= thalamus; TRIG= pontine trigeminal nuclei; BT = below positive threshold amplification.

| **Case** | **Sample** | **E Gene Ct** | **RNASE P Ct** |
| --- | --- | --- | --- |
| B1 | A17 | BT | 24 |
| B1 | AMYG | BT | 24 |
| B1 | CBL | BT | 23 |
| B1 | CP | BT | 24 |
| B1 | CSF | BT | 31 |
| B1 | DM | BT | 25 |
| B1 | ENT | BT | 24 |
| B1 | FC | BT | 24 |
| B1 | HIP | BT | 24 |
| B1 | HYT | BT | 24 |
| B1 | LEPTOS | BT | 24 |
| B1 | LN | BT | 24 |
| B1 | OB | BT | 24 |
| B1 | Serum | BT | 25 |
| B1 | SN | BT | 26 |
| B1 | TC | BT | 24 |
| B1 | THAL | BT | 25 |
| B1 | TRIG | BT | 25 |
| B2 | A17 | BT | 25 |
| B2 | AMYG | BT | 24 |
| B2 | CBL | BT | 23 |
| B2 | CP | BT | 24 |
| B2 | CSF | BT | 28 |
| B2 | DM | BT | 24 |
| B2 | ENT | BT | 24 |
| B2 | FC | BT | 24 |
| B2 | HIP | BT | 24 |
| B2 | HYT | BT | 25 |
| B2 | LEPTOS | BT | 24 |
| B2 | LN | BT | 24 |
| B2 | Lung | 35 | 24 |
| B2 | OB | BT | 24 |
| B2 | Serum | BT | 27 |
| B2 | SN | BT | 24 |
| B2 | TC | BT | 24 |
| B2 | THAL | BT | 25 |
| B2 | TRIG | BT | 25 |
| B3 | A17 | BT | 24 |
| B3 | AMYG | BT | 24 |
| B3 | CBL | BT | 23 |
| B3 | CP | BT | 25 |
| B3 | CSF | BT | 27 |
| B3 | DM | BT | 25 |
| B3 | ENT | BT | 24 |
| B3 | FC | BT | 23 |
| B3 | HIP | BT | 23 |
| B3 | HYT | BT | 23 |
| B3 | LEPTOS | BT | 24 |
| B3 | LN | BT | 24 |
| B3 | Lung | 30 | 24 |
| B3 | OB | BT | 24 |
| B3 | SN | BT | 24 |
| B3 | TC | BT | 23 |
| B3 | THAL | BT | 24 |
| B3 | TRIG | BT | 24 |
| B4 | A17 | BT | 26 |
| B4 | AMYG | BT | 24 |
| B4 | CBL | BT | 25 |
| B4 | CP | BT | 23 |
| B4 | CSF | BT | 32 |
| B4 | DM | BT | 25 |
| B4 | ENT | BT | 25 |
| B4 | FC | BT | 24 |
| B4 | HIP | BT | 24 |
| B4 | HYT | BT | 23 |
| B4 | LEPTOS | BT | 24 |
| B4 | LN | BT | 23 |
| B4 | Lung | 25 | 23 |
| B4 | OB | BT | 23 |
| B4 | SN | BT | 24 |
| B4 | TC | BT | 24 |
| B4 | THAL | BT | 25 |
| B4 | TRIG | BT | 32 |
| B5 | A17 | BT | 23 |
| B5 | AMYG | BT | 23 |
| B5 | CBL | BT | 23 |
| B5 | CP | BT | 24 |
| B5 | CSF | BT | 28 |
| B5 | DM | BT | 24 |
| B5 | ENT | BT | 24 |
| B5 | FC | BT | 24 |
| B5 | HIP | BT | 23 |
| B5 | HYT | BT | 23 |
| B5 | LEPTOS | BT | 24 |
| B5 | LN | BT | 23 |
| B5 | Lung | 28 | 24 |
| B5 | OB | BT | 24 |
| B5 | Serum | BT | 25 |
| B5 | SN | BT | 24 |
| B5 | TC | BT | 24 |
| B5 | THAL | BT | 25 |
| B5 | TRIG | BT | 25 |
| B6 | A17 | BT | 23 |
| B6 | AMYG | BT | 23 |
| B6 | CBL | BT | 23 |
| B6 | CP | BT | 24 |
| B6 | CSF | BT | 32 |
| B6 | ENT | BT | 24 |
| B6 | FC | BT | 23 |
| B6 | HIP | BT | 23 |
| B6 | HYT | BT | 22 |
| B6 | LEPTOS | BT | 24 |
| B6 | LN | BT | 23 |
| B6 | Lung | 35 | 24 |
| B6 | OB | BT | 24 |
| B6 | Serum | BT | 27 |
| B6 | SN | BT | 23 |
| B6 | TC | BT | 23 |
| B6 | THAL | BT | 24 |
| B7 | A17 | BT | 23 |
| B7 | AMYG | BT | 23 |
| B7 | CBL | BT | 22 |
| B7 | CP | BT | 24 |
| B7 | CSF | BT | 30 |
| B7 | DM | BT | 24 |
| B7 | ENT | BT | 23 |
| B7 | FC | BT | 23 |
| B7 | HIP | BT | 23 |
| B7 | HYT | BT | 22 |
| B7 | LEPTOS | BT | 24 |
| B7 | LN | BT | 23 |
| B7 | Lung | 31 | 23 |
| B7 | OB | BT | 24 |
| B7 | Serum | BT | 24 |
| B7 | SN | BT | 24 |
| B7 | TC | BT | 23 |
| B7 | THAL | BT | 24 |
| B7 | TRIG | BT | 24 |
| B8 | A17 | BT | 22 |
| B8 | AMYG | BT | 22 |
| B8 | CBL | BT | 22 |
| B8 | CP | BT | 23 |
| B8 | DM | BT | 21 |
| B8 | ENT | BT | 23 |
| B8 | FC | BT | 22 |
| B8 | HIP | BT | 23 |
| B8 | HYT | BT | 23 |
| B8 | Leptos | BT | 23 |
| B8 | LN | BT | 23 |
| B8 | Lung | BT | 22 |
| B8 | OB | BT | 24 |
| B8 | SN | BT | 23 |
| B8 | TC | BT | 22 |
| B8 | THAL | BT | 23 |
| B8 | TRIG | BT | 25 |
| B9 | A17 | BT | 24 |
| B9 | AMYG | BT | 23 |
| B9 | CBL | BT | 23 |
| B9 | CP | BT | 23 |
| B9 | DM | 40 | 24 |
| B9 | ENT | BT | 23 |
| B9 | FC | BT | 22 |
| B9 | HIP | BT | 24 |
| B9 | HYT | BT | 24 |
| B9 | LEPTOS | BT | 24 |
| B9 | LN | BT | 23 |
| B9 | Lung | 28 | 24 |
| B9 | OB | 28 | 23 |
| B9 | SN | BT | 27 |
| B9 | TC | BT | 23 |
| B9 | THAL | BT | 24 |
| B9 | TRIG | BT | 24 |
| B10 | A17 | BT | 23 |
| B10 | AMYG | 35 | 23 |
| B10 | CBL | BT | 23 |
| B10 | CP | BT | 23 |
| B10 | DM | BT | 24 |
| B10 | ENT | BT | 23 |
| B10 | FC | 34 | 23 |
| B10 | HIP | BT | 23 |
| B10 | HYT | BT | 23 |
| B10 | LEPTOS | BT | 23 |
| B10 | LN | BT | 23 |
| B10 | Lung | 18 | 24 |
| B10 | OB | BT | 27 |
| B10 | SN | BT | 23 |
| B10 | TC | 30 | 23 |
| B10 | THAL | BT | 23 |
| B10 | TRIG | BT | 23 |
| B11 | A17 | BT | 24 |
| B11 | AMYG | BT | 23 |
| B11 | CBL | BT | 23 |
| B11 | CP | BT | 24 |
| B11 | CSF | BT | 29 |
| B11 | DM | BT | 24 |
| B11 | ENT | BT | 23 |
| B11 | FC | BT | 23 |
| B11 | HIP | BT | 23 |
| B11 | HYT | BT | 22 |
| B11 | LEPTOS | BT | 24 |
| B11 | LN | BT | 23 |
| B11 | Lung | BT | 23 |
| B11 | OB | BT | 24 |
| B11 | Serum | BT | 26 |
| B11 | SN | BT | 23 |
| B11 | TC | BT | 23 |
| B11 | THAL | BT | 24 |
| B11 | TRIG | BT | 23 |
| B12 | A17 | BT | 23 |
| B12 | AMYG | BT | 23 |
| B12 | CBL | BT | 23 |
| B12 | CP | BT | 24 |
| B12 | CSF | BT | 34 |
| B12 | DM | BT | 24 |
| B12 | ENT | BT | 23 |
| B12 | FC | BT | 23 |
| B12 | HIP | BT | 23 |
| B12 | HYT | BT | 23 |
| B12 | LEPTOS | BT | 24 |
| B12 | LN | BT | 24 |
| B12 | Lung | BT | 24 |
| B12 | OB | BT | 24 |
| B12 | Serum | BT | 26 |
| B12 | SN | BT | 23 |
| B12 | TC | BT | 23 |
| B12 | THAL | BT | 23 |
| B12 | TRIG | BT | 23 |
| B13 | A17 | BT | 24 |
| B13 | AMYG | BT | 24 |
| B13 | CBL | BT | 23 |
| B13 | CP | BT | 23 |
| B13 | CSF | BT | 30 |
| B13 | DM | BT | 24 |
| B13 | ENT | BT | 25 |
| B13 | FC | BT | 24 |
| B13 | HIP | BT | 23 |
| B13 | HYT | 30 | 23 |
| B13 | LEPTOS | BT | 24 |
| B13 | LN | BT | 24 |
| B13 | Lung | BT | 24 |
| B13 | OB | BT | 24 |
| B13 | Serum | BT | 24 |
| B13 | SN | BT | 25 |
| B13 | TC | BT | 24 |
| B13 | THAL | BT | 27 |
| B13 | TRIG | BT | 25 |
| B14 | A17 | BT | 23 |
| B14 | AMYG | BT | 23 |
| B14 | CBL | BT | 24 |
| B14 | CP | BT | 24 |
| B14 | CSF | BT | 32 |
| B14 | DM | BT | 25 |
| B14 | ENT | BT | 24 |
| B14 | FC | BT | 23 |
| B14 | HIP | 25 | 23 |
| B14 | HYT | BT | 22 |
| B14 | LEPTOS | BT | 24 |
| B14 | LN | BT | 23 |
| B14 | Lung | BT | 24 |
| B14 | OB | BT | 24 |
| B14 | Serum | BT | 27 |
| B14 | SN | BT | 23 |
| B14 | TC | BT | 24 |
| B14 | THAL | BT | 24 |
| B14 | TRIG | BT | 25 |
| M1 | A17 | BT | 22 |
| M1 | AMYG | BT | 22 |
| M1 | CBL | BT | 22 |
| M1 | CP | BT | 23 |
| M1 | DM | BT | 21 |
| M1 | ENT | BT | 23 |
| M1 | FC | BT | 22 |
| M1 | HIP | BT | 23 |
| M1 | HYT | BT | 22 |
| M1 | LEPTOS | BT | 23 |
| M1 | LN | BT | 23 |
| M1 | OB | BT | 24 |
| M1 | SN | BT | 23 |
| M1 | TC | BT | 23 |
| M1 | THAL | BT | 25 |
| M1 | TRIG | BT | 23 |
| M2 | A17 | BT | 23 |
| M2 | AMYG | BT | 22 |
| M2 | CBL | BT | 21 |
| M2 | CP | BT | 29 |
| M2 | DM | BT | 21 |
| M2 | ENT | BT | 23 |
| M2 | FC | BT | 22 |
| M2 | HIP | BT | 25 |
| M2 | HYT | BT | 22 |
| M2 | LEPTOS | BT | 27 |
| M2 | LN | BT | 24 |
| M2 | OB | BT | 28 |
| M2 | SN | BT | 24 |
| M2 | TC | BT | 23 |
| M2 | THAL | BT | 23 |
| M2 | TRIG | BT | 24 |
| M3 | A17 | BT | 24 |
| M3 | AMYG | BT | 23 |
| M3 | CBL | BT | 22 |
| M3 | CP | BT | 27 |
| M3 | DM | BT | 23 |
| M3 | ENT | BT | 23 |
| M3 | FC | BT | 27 |
| M3 | HIP | BT | 23 |
| M3 | HYT | BT | 22 |
| M3 | LEPTOS | BT | 27 |
| M3 | LN | BT | 24 |
| M3 | OB | BT | 24 |
| M3 | SN | BT | 29 |
| M3 | TC | BT | 24 |
| M3 | THAL | BT | 28 |
| M3 | TRIG | BT | 25 |
| M4 | A17 | BT | 23 |
| M4 | AMYG | BT | 22 |
| M4 | CBL | BT | 22 |
| M4 | CP | BT | 22 |
| M4 | DM | BT | 22 |
| M4 | ENT | BT | 22 |
| M4 | FC | BT | 22 |
| M4 | HIP | BT | 24 |
| M4 | HYT | 30 | 22 |
| M4 | LEPTOS | BT | 23 |
| M4 | LN | BT | 23 |
| M4 | OB | BT | 24 |
| M4 | SN | BT | 22 |
| M4 | TC | BT | 23 |
| M4 | THAL | BT | 23 |
| M4 | TRIG | BT | 31 |
| M5 | A17 | BT | 28 |
| M5 | AMYG | BT | 23 |
| M5 | CBL | BT | 22 |
| M5 | CP | BT | 23 |
| M5 | DM | BT | 33 |
| M5 | ENT | BT | 23 |
| M5 | FC | BT | 22 |
| M5 | HIP | BT | 26 |
| M5 | HYT | BT | 23 |
| M5 | LEPTOS | BT | 24 |
| M5 | LN | 25 | 24 |
| M5 | SN | BT | 24 |
| M5 | TC | BT | 23 |
| M5 | THAL | BT | 31 |
| M5 | TRIG | BT | 30 |
| M6 | A17 | BT | 22 |
| M6 | AMYG | BT | 22 |
| M6 | CBL | BT | 22 |
| M6 | CP | BT | 22 |
| M6 | DM | BT | 20 |
| M6 | ENT | BT | 22 |
| M6 | FC | BT | 22 |
| M6 | HIP | BT | 26 |
| M6 | HYT | BT | 22 |
| M6 | LEPTOS | BT | 25 |
| M6 | LN | BT | 23 |
| M6 | OB | BT | 24 |
| M6 | SN | BT | 26 |
| M6 | TC | BT | 22 |
| M6 | THAL | BT | 25 |
| M6 | TRIG | BT | 24 |
| M7 | A17 | BT | 23 |
| M7 | AMYG | BT | 23 |
| M7 | CBL | BT | 22 |
| M7 | CP | BT | 23 |
| M7 | DM | BT | 21 |
| M7 | ENT | BT | 22 |
| M7 | FC | BT | 24 |
| M7 | HIP | BT | 24 |
| M7 | HYT | BT | 22 |
| M7 | LEPTOS | BT | 24 |
| M7 | LN | BT | 25 |
| M7 | OB | BT | 24 |
| M7 | SN | BT | 24 |
| M7 | TC | BT | 23 |
| M7 | THAL | BT | 23 |
| M7 | TRIG | BT | 25 |
| M8 | A17 | BT | 25 |
| M8 | AMYG | BT | 24 |
| M8 | CBL | BT | 23 |
| M8 | CP | BT | 24 |
| M8 | DM | BT | 24 |
| M8 | ENT | BT | 22 |
| M8 | FC | BT | 22 |
| M8 | HIP | BT | BT |
| M8 | HYT | BT | 22 |
| M8 | LEPTOS | BT | 28 |
| M8 | LN | BT | 24 |
| M8 | OB | BT | 25 |
| M8 | SN | BT | 28 |
| M8 | TC | BT | 23 |
| M8 | THAL | BT | 30 |
| M8 | TRIG | BT | 30 |
| M9 | A17 | BT | 21 |
| M9 | AMYG | BT | 23 |
| M9 | CBL | BT | 21 |
| M9 | CP | BT | 23 |
| M9 | DM | BT | 20 |
| M9 | ENT | BT | 22 |
| M9 | FC | BT | 21 |
| M9 | HIP | BT | 24 |
| M9 | HYT | BT | 22 |
| M9 | LEPTOS | BT | 24 |
| M9 | LN | BT | 23 |
| M9 | OB | BT | 23 |
| M9 | SN | BT | 24 |
| M9 | TC | BT | 22 |
| M9 | THAL | BT | 23 |
| M9 | TRIG | BT | 19 |
| M10 | A17 | BT | 23 |
| M10 | AMYG | BT | 23 |
| M10 | CBL | BT | 22 |
| M10 | CP | BT | 25 |
| M10 | DM | BT | 23 |
| M10 | ENT | 36 | 24 |
| M10 | FC | BT | 22 |
| M10 | HIP | BT | 28 |
| M10 | HYT | BT | 27 |
| M10 | LEPTOS | 34 | 24 |
| M10 | LN | BT | 28 |
| M10 | OB | BT | 25 |
| M10 | SN | BT | 23 |
| M10 | TC | BT | 23 |
| M10 | THAL | BT | 29 |
| M10 | TRIG | BT | 27 |
| M11 | A17 | BT | 22 |
| M11 | AMYG | BT | 22 |
| M11 | CBL | BT | 21 |
| M11 | CP | BT | 22 |
| M11 | DM | 25 | 20 |
| M11 | ENT | BT | 22 |
| M11 | FC | BT | 22 |
| M11 | HIP | BT | 23 |
| M11 | HYT | BT | 22 |
| M11 | LEPTOS | BT | 24 |
| M11 | LN | BT | 23 |
| M11 | OB | 29 | 19 |
| M11 | SN | BT | 28 |
| M11 | TC | BT | 22 |
| M11 | THAL | BT | 25 |
| M11 | TRIG | BT | 24 |
